# High incidence and geographic distribution of cleft palate cases in Finland are associated with a regulatory variant in *IRF6*

**DOI:** 10.1101/2024.07.09.24310146

**Authors:** Fedik Rahimov, Pekka Nieminen, Priyanka Kumari, Emma Juuri, Tiit Nikopensius, Kitt Paraiso, Jakob German, Antti Karvanen, Mart Kals, Abdelrahman G. Elnahas, Juha Karjalainen, Mitja Kurki, Aarno Palotie, FinnGen, Estonian Biobank Research Team, Arja Heliövaara, Tõnu Esko, Sakari Jukarainen, Priit Palta, Andrea Ganna, Anjali P. Patni, Daniel Mar, Karol Bomsztyk, Julie Mathieu, Hannele Ruohola-Baker, Axel Visel, Walid D. Fakhouri, Brian C. Schutte, Robert A. Cornell, David P. Rice

**Affiliations:** Department of Human Genetics, Genomics Research Center, AbbVie, Inc., North Chicago, IL 60064, USA; Department of Orthodontics, University of Helsinki, Helsinki 00014, Finland; Department of Anatomy and Cell Biology, University of Iowa, Iowa City, IA 52242, USA; Department of Oral Health Sciences, University of Washington, Seattle, WA 98195, USA; Department of Orthodontics, University of Helsinki and Helsinki University Hospital, Helsinki 00014, Finland; Estonian Genome Center, Institute of Genomics, University of Tartu, Tartu 51010, Estonia; Environmental Genomics and Systems Biology Division, Lawrence Berkeley Laboratories, Berkeley, CA 94720, USA; U.S. Department of Energy Joint Genome Institute, Lawrence Berkeley Laboratories, Berkeley, CA 94720, USA; Institute for Molecular Medicine Finland (FIMM), Helsinki Institute of Life Science (HiLIFE), University of Helsinki, Helsinki 00014, Finland; Analytic and Translational Genetics Unit, Massachusetts General Hospital, Boston, MA 02114, USA; Broad Institute of Harvard and MIT, Cambridge, MA 02142, USA; Cleft Palate and Craniofacial Center, Department of Plastic Surgery, University of Helsinki and Helsinki University Hospital, 00029 HUS, Finland; Department of Biochemistry, University of Washington School of Medicine, Seattle, WA 98195, USA; Institute for Stem Cell and Regenerative Medicine, University of Washington School of Medicine, Seattle, WA 98109, USA; Cancer Biology and Stem Cell Biology Laboratory, Department of Genetic Engineering, School of Bioengineering, College of Engineering and Technology, SRM Institute of Science and Technology, Chennai 603203, India; UW Medicine South Lake Union, University of Washington, Seattle, WA 98109, USA; Matchstick Technologies, Inc, Kirkland, WA 98033, USA; Department of Comparative Medicine, University of Washington School of Medicine, Seattle, WA 98195, USA; Brotman Baty Institute for Precision Medicine, Seattle, WA 98195, USA; Department of Bioengineering, University of Washington, Seattle, WA 98195, USA; School of Natural Sciences, University of California, Merced, CA 95343, USA; Department of Diagnostic and Biomedical Sciences, School of Dentistry, University of Texas Health Science Center at Houston, Houston, TX 77054, USA; Department of Pediatrics, McGovern Medical School, University of Texas Health Science Center at Houston, TX, 77030; Department of Microbiology and Molecular Genetics, College of Osteopathic Medicine, Michigan State University, East Lansing, MI 48824, USA

## Abstract

In Finland the frequency of isolated cleft palate (CP) is higher than that of isolated cleft lip with or without cleft palate (CL/P). This trend contrasts to that in other European countries but its genetic underpinnings are unknown. We performed a genome-wide association study for orofacial clefts, which include CL/P and CP, in the Finnish population. We identified rs570516915, a single nucleotide polymorphism that is highly enriched in Finns and Estonians, as being strongly associated with CP (*P* = 5.25 × 10^−34^, OR = 8.65, 95% CI 6.11–12.25), but not with CL/P (*P* = 7.2 × 10^−5^), with genome-wide significance. The risk allele frequency of rs570516915 parallels the regional variation of CP prevalence in Finland, and the association was replicated in independent cohorts of CP cases from Finland (*P* = 8.82 × 10^−28^) and Estonia (*P* = 1.25 × 10^−5^). The risk allele of rs570516915 disrupts a conserved binding site for the transcription factor IRF6 within a previously characterized enhancer upstream of the *IRF6* gene. Through reporter assay experiments we found that the risk allele of rs570516915 diminishes the enhancer activity. Oral epithelial cells derived from CRISPR-Cas9 edited induced pluripotent stem cells demonstrate that the CP-associated allele of rs570516915 concomitantly decreases the binding of IRF6 and the expression level of *IRF6*, suggesting impaired *IRF6* autoregulation as a molecular mechanism underlying the risk for CP.

## Introduction

Non-syndromic orofacial clefts (OFCs) are congenital malformations that affect approximately 1.7 per 1,000 newborns globally.^1^ The most common forms of OFCs can be classified as cleft lip only (CL), cleft lip with cleft palate (CLP) or cleft palate only (CP). Epidemiologically, CL and CLP are usually combined (CL/P, i.e., CL with or without CP) because it is unclear whether the cleft palate in individual cases of CLP is secondary to the cleft of the lip which occurs before secondary palate development. The incidence of specific OFCs varies based on the ethnicity and geographic origin of affected families.^1^ In non-Finnish European populations, the prevalence of CL/P is higher that of CP (0.80 per 1,000 births versus 0.55 per 1,000 births, respectively), as reported by EUROCAT^2,3^ for the years 1980–2020. However, Finland has one of the highest rates of non-syndromic (or isolated) CP in the world and the ratio of incidence of CL/P versus CP is reversed, with CP being more common.^4^ Remarkably, among the five Nordic countries where the total incidence of clefts is very similar, Finland stands out for having a higher CP incidence than CL/P; for instance, the CP incidence in Finland is 1.8-fold that in neighboring Sweden.^4^ In Finland the prevalence of births with CL/P over the period 2000–2014 was 1.08 per 1,000 births, whereas the prevalence of CP was 1.50 per 1,000 births.^5^ Furthermore, there is a significant regional variation in the incidence rate of CP, which increases from the lowest in the south-western-most to highest in the north-eastern-most province of the country.^5,6^ Moreover, 20% of OFC cases also have a family history of clefts, among whom the highest incidence (77%) was documented for cases with CP.^7^ These epidemiological observations suggest the presence of genetic risk factor(s) that contribute to the exceptionally high incidence and regional variation of CP rate in the Finnish population.^4^

Previous efforts to detect genetic risk factors that influence risk for CP include six genome-wide association studies (GWAS) which identified 15 loci associated with CP.^8–13^ Candidate genes at these loci are *GRHL3, IRF6*, *CTNNA2*, *POMGNT2*, *WHSC1*, *PARK2, MYC, PTCH1, YAP1, DOCK9*, *PAX9*, *DLK1*, *ISL2/SCAPER*, *FOXC2*-*FOXL1* and *MAU2*. In one study, variants near *MLLT3* and *SMC2* were associated with increased risk for CP but only if the mother consumed alcohol during the peri-conceptual period, as were variants near *TBK1* and *ZNF236* but only in the presence of maternal smoking.^12^

At just one locus, 1p36, the causative variant has been identified and, unusually, it is a coding variant.^13,14^ This variant is a missense single nucleotide polymorphism (SNP) in the *GRHL3* gene that disrupts the transactivation function of the transcription factor encoded by this gene. GRHL3 is important for keratinocyte and periderm differentiation^15–17^, as is IRF6^18,19^, and the *IRF6* gene is also associated with CP.^8,9,11^ In zebrafish, Grhl3 acts downstream of Irf6 in periderm differentiation.^20^ Together these findings imply the perturbation of a gene regulatory network involving IRF6 and GRHL3, which orchestrates the differentiation of oral periderm, predisposes individuals to CP. Abnormal periderm differentiation is also proposed to underlie some cases of CL/P.^21^ However, while variants associated with non-syndromic CL/P have been detected near *IRF6*, none have thus far been reported near *GRHL3*. Indeed, among hundreds of genes connected to orofacial clefts, *IRF6* is one of only two genes (*MSX1*), to date, in which rare mutations cause syndromic forms of both CL/P and CP, and common DNA variants contribute risk for non-syndromic forms of both CL/P and CP.

Here we conducted a GWAS in the Finnish population and report a novel association between isolated CP and the *IRF6* gene locus. The risk allele is a low-frequency, non-coding SNP located in MCS-9.7, a known enhancer for the *IRF6* gene.^22^ Using *in vitro* assays, we find evidence that the variant directly affects *IRF6* expression through altered binding of IRF6 to this enhancer. This study adds to the short list of common variants whose contribution to etiology of non-syndromic OFCs has been evaluated in molecular detail. To our knowledge this is the first example of variants in a single enhancer being associated with risk for both CP and with CL/P.

## Materials and methods

### FinnGen cohorts

The FinnGen study is a population-based cohort study launched in 2017 as a public-private partnership aiming to identify genotype-phenotype relationships in the Finnish founder population.^23^ Three national and six regional biobanks provide samples to the FinnGen study. Individuals from several previously established prospective epidemiological and disease cohort studies are also included in FinnGen. Participants in FinnGen provided informed consent for biobank research in accordance with the Finnish Biobank Act (law 688/2012). Alternatively, separate research cohorts, collected prior to when the Finnish Biobank Act came into effect (in September 2013) and the start of FinnGen (August 2017), were collected based on study-specific consents and later transferred to the Finnish biobanks after approval by Fimea (Finnish Medicines Agency), the National Supervisory Authority for Welfare and Health. Recruitment protocols followed the biobank protocols approved by Fimea. Demographic and clinical data are derived from nationwide electronic health registers. These registers contain records of major health-related events, such as hospitalizations, prescription drug purchases, medical procedures, or deaths with a data collection history spanning over 50 years. This study included 309,154 participants from Data Freeze (DF) 7 as the discovery cohort and 200,100 participants from DF8 through DF12 as the replication cohort. All participants had a Finnish genetic ancestry determined by a principal component (PC) analysis of their genotypes.

### Estonian cohort

Estonian Biobank is a population-based biobank at the Institute of Genomics, University of Tartu. The current cohort size is 200,000 individuals (aged ≥ 18), reflecting the ethnicities, ages, sex, and geographical distribution of the adult Estonian population. All subjects have been recruited by general practitioners and physicians in hospitals and during promotional events. Upon recruitment, all participants completed a questionnaire about their health status, lifestyle, and diet. The Estonian Biobank database is linked with national registries, such as Cancer Registry and Causes of Death Registry, hospital databases, and the national health insurance fund database, which holds treatment and procedure service bills. Diseases and health-related conditions are recorded as International Classification of Diseases (ICD) codes (version 10). These health data are continuously updated through periodical linking to national electronic databases and registries. An in-depth description of Estonian Biobank can be found elsewhere.^24,25^ Ethical approval for this study was obtained from the Research Ethics Review Committee of the University of Tartu. All participants signed informed consent.

### Phenotype definitions

Case subjects with non-syndromic forms of OFCs in FinnGen were defined from electronic health records in the Care Register for Health Care of the Finnish Institute for Health and Welfare (THL) and the Causes of Death Register from Statistics Finland, using diagnosis codes of the Finnish version of ICD (versions 10, 9, 8). Codes of versions ICD-10, ICD-9 and ICD-8 were available since 1996, between 1987-1995 and between1967-1986, respectively. Cases with CP only (discovery cohort n = 228; 157 females and 71 males and replication cohort n = 165; 116 females and 49 males) were defined based on the ICD-10 code Q35, ICD-9 code 7490[ABCE], and ICD-8 code 74900. Cases with CLP (n = 121; 62 females and 59 males) were defined using the ICD-10 code Q37, ICD9 code 7492[ABCDE], and ICD8 codes 74920 and 74924. Cases with CL (n = 54; 28 females and 26 males) were defined using ICD10 code Q36, ICD9 codes 7491[ABCDEFG] and ICD8 codes 74910 and 7491[123]. Syndromic cases with the following ICD-10 codes were excluded: Q75.4 (mandibulofacial dysostosis), Q38.0 (congenital malformations of lips, not elsewhere classified, including VWS), Q86 (congenital malformation syndromes due to known exogenous causes, not elsewhere classified), Q87 (other specified congenital malformation syndromes affecting multiple systems) and Q91 (Edward’s syndrome and Patau’s syndrome). The Estonian cohort consisted of 71 CP cases and 198,973 population controls. Cases with isolated CP (n = 71; 54 females and 17 males) in the Estonian biobank were ascertained using the ICD-10 code Q35 through a questionnaire implemented by the biobank and national health registry-based medical records. The following ICD-10 codes for syndromic cases were excluded among Estonian cases and controls: Q38.0 (congenital malformations of lips, not elsewhere classified, including VWS), Q38.1 (ankyloglossia), Q38.5 (congenital malformation of palate), Q87 (other specified congenital malformation syndromes affecting multiple systems), and Q91 (Edwards syndrome and Patau syndrome).

### Genotyping and imputation

Samples collected and genotyped before the launch of the FinnGen study were genotyped with a mix of Illumina (Illumina) and Affymetrix arrays (Thermo Fisher Scientific). However, recently collected samples were genotyped on two versions of a custom designed Affymetrix array, referred to as FinnGen1 (657,675 markers) and FinnGen2 (664,510 markers), with 655,973 overlapping markers. Array content, sample genotyping and calling, imputation, PC analysis and estimation of population structure and ancestry, and quality control (QC) procedures are described in more detail by Kurki et al.^23^ Briefly, genotype calls were made with GenCall and zCall algorithms for Illumina, and AxiomGT1 algorithm for Affymetrix data. Data generated on previous reference genome builds were lifted over to build version 38 (GRCh38/hg38), as described in https://dx.doi.org/10.17504/protocols.io.xbhfij6. After basic QC, such as exclusion of samples with missing sex information and those that were mixed up, sample and variant QCs were performed as follows. In sample-wise QC, individuals whose genetic sex mismatched sex provided from registries, with high genotype missingness (>5%), excess heterozygosity (±4SD) and non-Finnish ancestry were excluded. In variant-wise QC, variants with high missingness (>2%), deviation from Hardy–Weinberg equilibrium (*P* < 1 × 10^−6^) and minor allele count (MAC) < 3 were excluded. Samples passing QC were then pre-phased chip-wise with Eagle (v2.3.5)^26^ using the default parameters, except that the number of conditioning haplotypes was set to 20,000. Finally, genotype imputation was carried out with Beagle 4.1 (version 08Jun17.d8b)^27^ by using a Finnish population-specific reference panel built from 3,775 high-coverage (25-30×) whole-genome sequences (WGS) in Finns, as described in the following protocol: https://dx.doi.org/10.17504/protocols.io.nmndc5e. Approximately 17M variants were imputed. Variants with an imputation INFO score < 0.8 and minor allele frequency (MAF) < 0.0001 were excluded, resulting in a combination of 15.36M directly genotyped and imputed variants included in association tests. In total, 309,154 individuals (173,746 females and 135,408 males) passed QC and were included in the discovery GWAS, and 200,100 individuals (113,132 females and 86,968 males) were included in the replication GWAS. Samples from the Estonian Biobank cohort were genotyped at the Core Genotyping Lab of the Institute of Genomics, University of Tartu, using Illumina Infinium Global Screening arrays v1.0, v2.0 and v2.0_EST. Individuals whose sex defined based on chromosome X heterozygosity did not match the sex recorded in phenotype data were excluded from the analysis. Before pre-phasing and genotype imputation, genotyped variants were filtered by call rate <95%, Hardy–Weinberg equilibrium *P* < 1 × 10^−4^ (autosomal variants only), and minor allele frequency of 1%. Pre-phasing was performed using Eagle (v2.3.5)^26^, and imputation was performed using Beagle 5.0 (v.28Sep18.793)^27^ by using an Estonian population-specific imputation reference panel consisting of 2,297 high-coverage (∼30×) WGS data from Estonian individuals.^28^

### Association analysis

Association analyses were carried out using the REGENIE program, which uses a two-step, whole-genome regression framework that controls for population stratification and sample relatedness.^29^ We used the code version 2.0.2 of the program which was modified to include dosage-based calculation of allele frequencies in cases and controls. For step 1 LOCO prediction, we included sex as determined from genotypes, age, genotyping batch, and the first 10 PCs as covariates. We used a genotype block size of 1,000. Genetic relatedness in step 1 was calculated using a set of 55,139 linkage disequilibrium (LD)-pruned, well-imputed, high-quality (INFO > 0.95 and MAF > 0.01) SNPs. LD pruning was performed with PLINK (v2.00a2.3LM)^30^ with a window size of 1 Mb and *r*^2^ = 0.1. Association of the phenotypes with all imputed genetic markers with INFO ≥ 0.8 and MAF ≥ 0.0001 were tested in step 2 using the logistic regression score test. To better control the type I error for rare and low-frequency genetic markers due to severely imbalanced case-control ratios, we used the approximate Firth test for variants with an initial *P* value of less than 0.01 and computed the standard error based on effect size and likelihood ratio test *P* value (regenie options *--firth --approx --pThresh* 0.01 *--firth-se*). Association testing in the Estonian Biobank cohort data was also performed using REGENIE (v2.2.4), including sex, age and the first 10 PCs as covariates. A minimum MAC of 5 was used when testing variants. Similar to the FinnGen GWAS, the approximate Firth test was applied to control type I error for rare and low-frequency variants due to severely imbalanced case-control ratios, with the following parameters (regenie options *--firth --approx --pThresh* 0.01 *--firth-se*). Manhattan plots were generated using the *ggplot2* R package.^31^ Regional association plots were generated with the stand-alone version of LocusZoom (v1.3),^32^ using the Finnish population-specific LD structure estimated in 3,775 whole-genome sequences of Finns. To test genotype distributions, we used BCFtools^33^ (v1.18) to extract SNP genotypes from VCF files, counted the number of genotypes in R (v4.3.2) and used a chi-squared test for independence of distributions of cases and controls between genotypes.

### Meta-analysis

The lead SNPs identified in discovery GWAS were meta-analyzed using FinnGen replication and Estonian Biobank GWAS summary statistics based on the inverse-variance weighted fixed effects model with the METAL software^34^ (version 2011-03-25).

### Conditional analysis

Conditional analysis was performed using the COJO^35^ approach (*--cojo-cond* command) implemented in the GCTA software (v1.93.2).^36^ For this analysis, we used the summary statistics output from the REGENIE GWAS and imputed genotypes from the entire FinnGen cohort for LD estimation within 1 Mb in each direction from the conditioning SNP.

### FinRegistry

To assess the geographic distribution of non-syndromic CP prevalence in different regions of Finland, we used data from a nationwide cohort (FinRegistry, https://www.finregistry.fi), including 5,216,731 individuals. We assigned each individual to one of the 19 administrative regions based on their first recorded address. CP cases were defined using the same ICD code-based inclusion and exclusion criteria as in FinnGen (see Phenotype definitions). The prevalence was estimated by dividing the number of CP cases in each region by the total number of individuals in that region.

### Geographic variation

The geographic distribution of allele frequencies was calculated and plotted using region-level, birthplace data of 306,678 FinnGen study participants obtained from Statistics Finland, with the *ggplot2*, *ggrepel* and *sf*^37^ R packages. Spatial polygon data (v3.6) for the Finnish map were downloaded from the database of Global Administrative Areas. Correlation between the regional prevalence of CP and allele frequency was calculated and plotted using Pearson’s product-moment correlation in base R (v3.6.1).

### Population attributable risk

The population attributable risk (PAR) is the proportion of cases that would be prevented if a risk factor were eliminated from the population. Individual and combined PAR of risk variants were calculated as described previously, using the following formulas:^38^

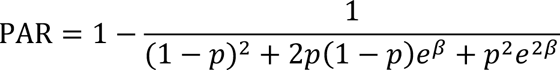

where *p* is the population frequency of the risk allele and *β* is the beta coefficient from the regression analysis, which shows the effect size of the variant on the phenotype and *e^β^* expresses odds ratio (OR), and

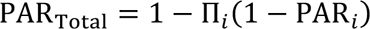

where the risk variants are assumed to be independent of one another and that their combined effects on disease are multiplicative.

### Bioinformatic analysis

SNP overlap with open-chromatin peaks, chromatin immunoprecipitation-sequencing (ChIP-Seq) peaks of histone activation marker (H3K27ac) from HIOEC oral epithelial cells and publicly available chromatin modification datasets from 9 principal cell types from the ENCODE Project Consortium^39^ and from different stages of embryonic facial tissues from the Cotney lab^40^ were visualized using the UCSC Genome Browser.

H3K27Ac ChIP-Seq data from HIOEC oral epithelial cells and ATAC-Seq (Assay for Transposase-Accessible Chromatin using sequencing) data from both the HIOEC oral epithelial cells and the HEPM oral mesenchymal cells were generated previously.^41^ *In silico* transcription factor binding activities at the risk variant site were determined using HOMER^42^ and the Transcription factor Affinity Prediction (TRAP) tool.^43^ Position Weight Matrix of the consensus IRF6 binding motif was obtained from the JASPAR database.^44^

### Cell lines

Primary Epidermal Keratinocytes: Normal, Human, Neonatal Foreskin (HEKn) purchased from ATCC (ATCC PCS-200-010) were maintained and cultured as per manufacturer’s instruction in dermal cell basal medium (ATCC PCS-200-030) supplemented with keratinocyte growth kit (ATCC PCS-200-040). The human induced pluripotent stem cells (iPSCs), WTC-11 (Coriell Institute for Medical Research, Catalog no. GM25256), were maintained in mTeSR1 media (StemCell Technologies) on Matrigel (Corning) coated plates and passaged with Accutase. Cells were incubated at 37°C in 5% CO_2_.

### Electroporation and dual luciferase reporter assay

Each firefly reporter construct (Supplemental Notes) was co-transfected with a constitutively driven Renilla luciferase plasmid for dual luciferase assays. Briefly, HEKn cells were electroporated using the Amaxa Cell Line Nucleofector Kit (Lonza) and the Nucleofector II instrument (Lonza) program T-025. We used a dual-luciferase reporter assay system (Promega) and FB12 Luminometer (Berthold Detection Systems) to evaluate the luciferase activity following the manufacturer’s instructions. Relative luciferase activity was calculated as the normalized values of the Firefly to the Renilla enzyme activities. Three independent measurements were performed for each transfection group, and three biological replicates were performed. All results were presented as mean ± standard deviation (s. d.). Statistical significance was determined using the Student’s *t*-test (two-tailed).

### Transgenic mouse reporter assays

MCS-9.7*-lacZ* transgenic reporter constructs were generated with the risk and non-risk alleles of rs570516915 on the Finnish haplotype background with a mouse *Shh* promoter (Supplementary Notes). These constructs were co-injected with the Cas9 protein and gRNA targeting the H11 locus^45^ into the mouse embryos, and embryos were harvested at E13.5 and processed for X-gal staining as described previously.^22^ Transgenic mouse embryos were imaged with Adobe Photoshop Elements 11. Plasmid copy number integration for transgenic embryos was estimated by qPCR with probes targeting the mouse *Shh* promoter region (Mm00560391_cn, Thermo Fisher Catalog no. 4400291). This probe targets both endogenous and plasmid copies. The *Tfrc* gene was used as a reference locus (Thermo Fisher Catalog no. 4458366). Three non-transgenic mouse DNA samples were used as normalization controls. To calculate an estimated plasmid integration copy number, the 2^−ΔΔ*Ct*^ approach was used. Stained embryos were visually assessed and embryos with no staining at all were excluded from the analysis. Embryos were ordered according to the staining intensities in the heads, from the strongest to the weakest, by 14 (2 batches) and 7 (1 batch) independent observers who were blinded to the genotype of the reporter construct.^45^ In each assessment, the embryos were given a score based to their position in the order (1, 2, etc.) and sums of these were used to define a consensus order (Supplementary Fig. 1). Mean positions were calculated for the non-risk and risk allele carrying embryos in each assessment and means of these mean values (Supplementary Table S1) were then compared using the two-tailed Wilcoxon Signed Ranks test with the IBM SPSS Statistics software (v29.0.0.0). Mice were housed in the animal facility where their health, food, water, and housing conditions were monitored with daily visual checks by technicians. Mice were housed in bioBUBBLE Clean Rooms, soft-walled enclosures powered by 80-100 air changes per hour of HEPA filtration under Light/Dark Cycle of 12:12 starting at 6AM, at 20.5-24.4°C, and humidity 30-70%. Mouse experiments were performed at the Lawrence Berkeley National Laboratory (LBNL) and were reviewed and approved by the LBNL Animal Welfare and Research Committee.

### CRISPR-Cas9 mediated genome editing

Integrated DNA Technologies (IDT) CRISPR-Cas9 guide RNA (gRNA) design checker tool was used to design specific CRISPR RNA (crRNA) harboring the rs570516915 in the seed region to avoid the re-cleavage by CRISPR-Cas9 (Supplementary Fig. 2). A homology directed repair (HDR) template including the desired mutation was designed to have a minimum of 30-35nt long homology arms (Supplementary Fig. 2). Briefly, equimolar concentrations of crRNA and trans-activating crRNA (tracrRNA, IDT: 1072532) were annealed at 95°C for 5 min followed by cooling at room temperature (RT) to form functional gRNA duplexes. Further, the ribonucleoprotein (RNP) complex was prepared 15 minutes before transfection by mixing gRNAs (1.5µM) with Cas9 protein (0.45µM) (Sigma, Catalog no. CAS9PROT) at RT. RNP complexes together with the repair template were transfected into 1 million WTC11-iPSCs with Amaxa nucleofector (Lonza, Human Stem Cell kit 2, program: A-023) in presence of ROCK inhibitor (Y-27632, Catalog no. 04-0012-10, Stemgent; [10mM] stock). Individual colonies were hand-picked and plated into 96 well plates. DNA was extracted using Quick Extract DNA extraction solution (Epicentre Catalog no. QE09050). Colonies were screened by PCR and sequenced using primers flanking rs570516915 (Primer sequences are provided in Supplementary Table 2). Three independent clones each of heterozygous (TG), and homozygous risk (GG), and two clones of the homozygous non-risk (TT) genotype were isolated.

### *In vitro* differentiation of iPSCs into embryonic oral epithelial cells

Three independent samples of each genotype (3 clones of the heterozygous genotype, 3 clones of the homozygous risk genotype, and 2 clones plus one sample of the parental WTC-11 cells of the homozygous non-risk genotype) were seeded in triplicate in 12-well plate (50,000 cells per well). Further, the cells were differentiated to embryonic oral epithelial cells using a 10-day differentiation protocol described previously^46^ (Supplementary Fig. 3).

### Quantitative RT-PCR on induced embryonic oral epithelial cells

From nine samples of each genotype (Supplementary Fig. 3) RNA was extracted using Quick-DNA/RNA Miniprep kit (Zymo Research) followed by DNase treatment using manufacturer’s instructions (Thermo Fisher Scientific). Reverse transcription was performed using High-Capacity cDNA Reverse Transcription Kit (Thermo Fisher Scientific). Real-time PCR was performed using SYBR Green qPCR mix (Bio-Rad) in Bio-Rad CFX Connect Real-Time System. Quantitative RT-PCR reaction for each sample was performed in triplicate. *IRF6* expression level was normalized against five genes: four housekeeping genes (*ACTB*, *GAPDH*, *HPRT*, & *UBC*) and an epithelial marker, *CDH1*, whose expression is IRF6-independent^47^ using a robust method^48^. Data were presented as mean ± s. d. Statistical significance was determined using the Student’s *t*-test (two-tailed). Primer sequences used in the study for qRT-PCR are provided in Supplementary Table 2.

### Chromatin immunoprecipitation qPCR

Induced oral epithelial cells (iOECs) derived from iPSC edited to be heterozygous for risk and non-risk alleles of rs570516915 (three heterozygous clones differentiated to oral epithelial cells) were harvested (∼3.5 million cells) and fixed in 1% formaldehyde for 15 min at RT. Fixation was stopped with 125 mM glycine for 5 min at RT. Chromatin immunoprecipitation on iOECs was performed using a semi-automated protocol described previously.^49^ Briefly, cells were sheared using PIXUL to obtain genomic DNA fragments averaging 300 to 600 bp. The sonicated cell lysates were subjected to chromatin immunoprecipitation with specific antibody (1 μg of anti-H3K27Ac from Millipore-Sigma and 1 μg of Rabbit anti-IRF6 from Dr. Akira Kinoshita, Nagasaki University, as previously described).^50^ Normal rabbit IgG (1 μg) was used as negative antibody control. Obtained DNA was quantitated using Qubit dsDNA HS assay kit (Thermo Fisher Scientific). Quantitative PCR was performed with equal mass of DNA (1.25ng) in triplicate from three ChIP replicates of each antibody and percent input DNA was calculated using the method described previously.^51^ Primer sequences are shown in Supplementary Table 3. Briefly, Ct values obtained were normalized with input DNA and expressed as fold DNA enrichment over IgG. Negative control PCR primers were designed to target a sequence 103.7 kb upstream of the *IRF6* transcription start site that did not harbor active regulatory elements identified from ATAC-Seq and H3K27Ac ChIP-Seq in HIOEC or NHEK cells and devoid of predicted IRF6 binding sites. Data were presented as mean ± s. d. Statistical significance was determined using the Student’s *t*-test (two-tailed). GraphPad Prism 10 was used for the statistical data analysis for luciferase reporter assays, RNA expression and ChIP-qPCR experiments.

### Immunostaining on iPSCs and induced oral epithelial cells

Human iPSCs and iOECs were fixed in 4% paraformaldehyde (PFA) (EMS, Catalog no. 15710) for 12min at RT and later washed thrice with 1X PBS for 5 min each on a rocker followed by permeabilization with 0.5% TritonX-100 (Sigma, Catalog no. T9284) at RT on a rocker for 10 minutes. Later blocking was done for an hour at RT on a rocker with a blocking buffer consisting of 5% goat serum (VWR, Catalog no. 101098-382), 3% BSA (VWR, Catalog no. 9048-46-8), and 0.1% Triton X-100. Then, the cells were incubated overnight in the primary antibodies (PITX2, Santa Cruz, Catalog no. sc-390457, 1:50; ECAD/CDH1, ABclonal, Catalog no. A20798, 1:200) at 4°C on a rocker. Further, cells were rinsed with 1X PBS thrice for 5mins each and later incubated with corresponding secondary antibodies (Mouse IgG Alexa Flour 488, Thermo Fisher, Catalog no. A-11001, 1:500; Rabbit IgG Alexa Flour 568, Thermo Fisher, Catalog no. A-11036, 1:500) followed by Phalloidin (Phalloidin-iFlour 647 Reagent, abcam, Catalog no. ab176759, 1:500) for an hour at RT on a rocker prepared in the same blocking agent, followed by washing with 1X PBS thrice for 5min each in the dark. The cells were incubated in autofluorescence quenching solution (Vector Labs, Catalog no. SP-8400) for 5 min at RT under dark conditions on a rocker and rinsed with 1x with PBS. Cells were further incubated in DAPI (Thermo Fisher, Catalog no. D1306) for 10 min then rinsed with 1X PBS, mounted with Vectashield (Vector Labs, Catalog no. H-1700), and imaged using Leica TCS-SPE Confocal microscope. Images were processed with Fiji software distribution of ImageJ v2.9.0.

## Results

### A low-frequency SNP in the *IRF6* enhancer MCS-9.7 is associated with CP

We conducted a GWAS of non-syndromic OFC in the Finnish population using the FinnGen study cohort with a mixture of 355 CL/P and CP cases, and 308,799 ethnicity-matched population controls (Data Freeze 7). We identified genome-wide significant (*P* < 5 × 10^−8^) associations with OFC and two genomic loci located on the long and short arms of chromosome 1 (Supplementary Fig. 4a). Upon dividing the case group into subtypes of clefts, we discovered that the association signals at both loci were driven by the subgroup of 228 cases with CP (Fig. 1). GWAS of 151 cases with CL/P did not yield any genome-wide significant hits (Supplementary Fig. 4b). Due to small sample sizes, we did not conduct independent analyses in the CL and CLP subsets. Variants reaching *P* values less than 1.0 × 10^−4^ in all three association analyses are provided in Supplementary Tables 4, 5 and 6.

**Fig. 1.**
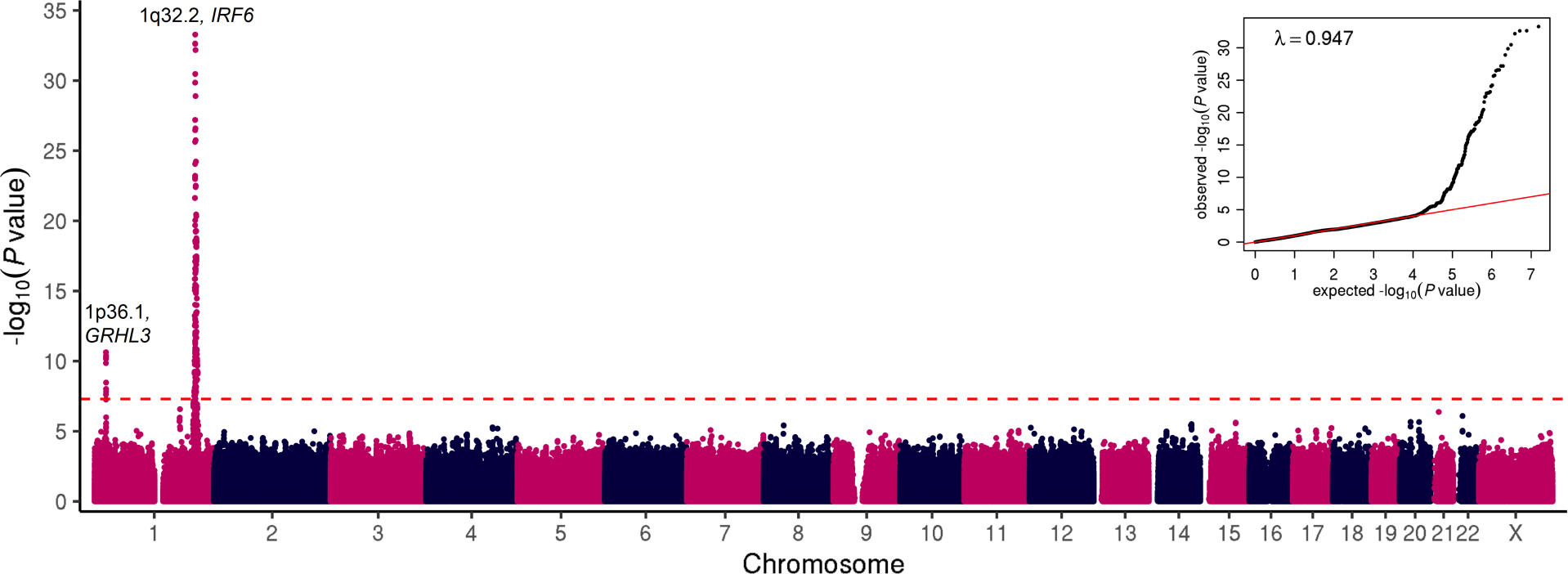
Manhattan and quantile-quantile (Q-Q) plots showing GWAS results of 228 cases affected with non-syndromic CP and 308,799 population controls in the FinnGen discovery cohort. Negative log_10_ *P* values are plotted for each variant against their chromosomal coordinates provided in the human genome build GRCh38/hg38. Two-sided *P* values were estimated using Firth regression and not adjusted for multiple hypothesis testing. Red dashed line represents the threshold for genome-wide significance (*P* = 5 × 10^−8^) after Bonferroni correction for multiple testing. A Q-Q plot is shown in the inset panel, where the observed *P* values are plotted against the expected *P* values under null distribution (red line).

In the GWAS of CP, ten SNPs reached genome-wide significance on chromosome 1p36.1, including the lead SNP rs113965554 (*P* = 2.32 × 10^−11^, OR = 2.73) and the previously described etiologic missense variant p.Thr454Met (rs41268753, *P* = 1.39 × 10^−10^, OR = 2.66) in the *GRHL3* gene (Table 1).^13,14^ The top five variants with similar *P* values are in strong LD with one another (Fig. 2a). These results indicate that the 1p36.1 signal is likely driven by the *GRHL3* missense variant, further replicating the association between this common missense SNP and CP in the Finnish population.^13^

**Fig. 2.**
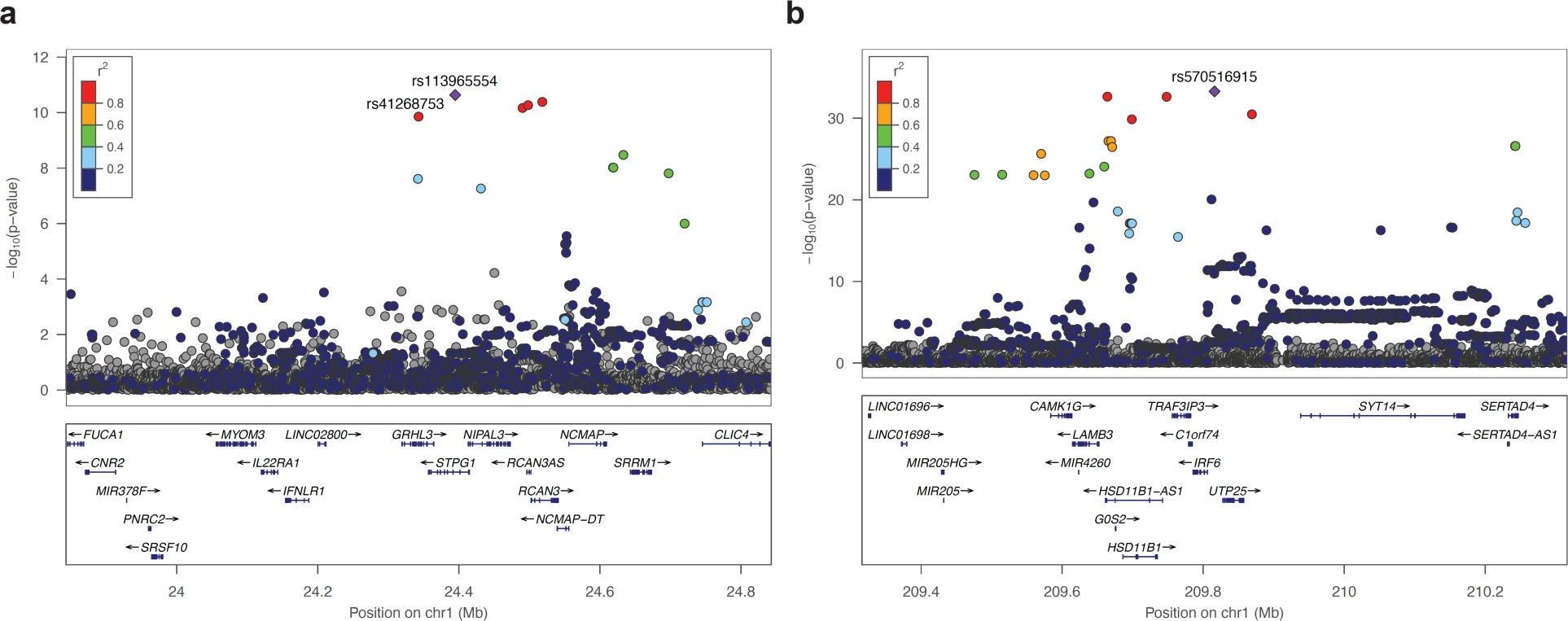
Regional association plots of the chromosome 1 risk loci for non-syndromic CP. Data are shown for association on chromosome (**a**) 1p36.1 (*GRHL3*) and (**b**) 1q32.2 (*IRF6*). Negative log_10_ *P* values (two-sided) are shown for variants within a 1Mb region centered at the reference SNP. Plotted *P* values are two-sided raw *P* values estimated using Firth regression. The reference SNP is marked with a purple diamond, and pairwise LD (*r*^2^) between the reference SNP and other variants are indicated by color. The *r*^2^ values were estimated from high-coverage whole-genome sequences of 3,775 Finns. Both directly genotyped and imputed SNPs are plotted. Genomic coordinates are shown according to the human genome build GRCh38/hg38.

**Table 1.** Discovery, replication, and meta-analysis of lead SNPs on chromosome 1p36.1 and 1q32.2 associated with CP in the Finnish and Estonian populations.

| SNP | RA | FinnGen Discovery<br>(228 cases, 308,799 controls) |  |  |  | FinnGen Replication<br>(165 cases, 199,935 controls) |  |  |  | Estonian Biobank<br>(71 cases, 198,973 controls) |  |  |  | Meta-analysis |  |  | RAF in gnomAD<br>(v3.1.2) |  |  |
| --- | --- | --- | --- | --- | --- | --- | --- | --- | --- | --- | --- | --- | --- | --- | --- | --- | --- | --- | --- |
|  |  | RAF | OR | 95% CI | P value | RAF | OR | 95% CI | P value | RAF | OR | 95% CI | P value | OR | 95% CI | P value | FIN | NFE | All |
| rs113965554*<br>(rs41268753) | A<br>(T) | 4.92<br>(4.89) | 2.73<br>(2.66) | 2.03–3.66<br>(1.97–3.58) | $2.32 \times 10^{-11}$<br>($1.39 \times 10^{-10}$) | 4.96<br>(4.91) | 2.15<br>(2.17) | 1.47–3.14<br>(1.49–3.17) | $7.37 \times 10^{-5}$<br>($5.64 \times 10^{-5}$) | 3.21<br>(3.21) | 2.39<br>(2.36) | 1.23–4.66<br>(1.21–4.57) | 0.01<br>(0.01) | 2.48<br>(2.45) | 1.99–3.09<br>(1.96–3.05) | $4.55 \times 10^{-16}$<br>($1.95 \times 10^{-15}$) | 5.19<br>(5.13) | 3.49<br>(3.37) | 2.35<br>(2.24) |
| rs7546104 | A | 24.11 | 1.95 | 1.64–2.33 | $9.53 \times 10^{-14}$ | 24.40 | 1.92 | 1.56–2.36 | $5.87 \times 10^{-10}$ | 23.69 | 1.83 | 1.32–2.55 | 0.0003 | 1.92 | 1.70–2.18 | $4.97 \times 10^{-25}$ | 25.01 | 17.50 | 13.39 |
| rs570516915 | G | 1.20 | 8.65 | 6.11–12.25 | $5.25 \times 10^{-34}$ | 1.29 | 9.39 | 6.28–14.04 | $8.82 \times 10^{-28}$ | 0.24 | 13.55 | 4.21–43.63 | $1.25 \times 10^{-5}$ | 9.14 | 8.89–9.40 | $4.34 \times 10^{-64}$ | 1.55 | 0.08 | 0.15** |

Two-sided *P* values in the FinnGen and Estonian Biobank cohorts were estimated using Firth regression and not adjusted for multiple hypothesis testing. *P* values of the meta-analysis are two-sided *P* values derived from a fixed-effect meta-analysis and were not adjusted for multiple testing.
\*rs113965554 and rs41268753 (the etiologic missense variant p.Thr454Met in *GRHL3*) are in nearly complete LD ( $r^2 = 0.99$ ) with each other.
\*\*Besides Finnish and non-Finnish Europeans, only 3 alleles were found in African/African Americans in the gnomAD (v3.1.2) database. RA – risk allele; OR – odds ratio; CI – confidence interval; RAF – risk allele frequency (%); FIN – Finnish; NFE – Non-Finnish European.

Most importantly, we observed significant association signals on chromosome 1q32.2, where a total of 240 variants spanning over 7 Mb reached genome-wide significance. Examination of allele frequencies and LD patterns suggested multiple independent signals in the region covering the *IRF6* gene (Fig. 2b). One set of variants comprised a 62 kb-long haplotype of strongly correlated variants upstream of *IRF6*, with the lead SNP rs7546104 reaching a *P* value of 9.53 × 10^−14^ (OR = 1.95, Table 1, Supplementary Fig. 5a-b). One of the SNPs within this haplotype, rs72741048 (*P* = 1.15 × 10^−11^, Supplementary Fig. 5b), has previously been linked to OFC, including CP, in a large GWAS in the Han Chinese population.^8^

A second, highly significant set of associated variants in 1q32.2 was defined by the lead SNP rs570516915 (*P* = 5.25 × 10^−34^, OR = 8.65, Fig. 2b). This signal is a novel association with CP and the lead SNP changes a highly-conserved nucleotide (Supplementary Fig. 6) in a previously-described *cis*-regulatory element 9.7 kb upstream of *IRF6* transcription start site (i.e., <u>m</u>ulti-species <u>c</u>onserved <u>s</u>equence, 9.7 kb, “MCS-9.7”).^22^ The low-frequency risk allele G (1.2%) of rs570516915 appears to have originated on the more prevalent haplotype tagged by the risk allele A (24.1%) of rs7546104. Subsequently, the rs570516915 variant has increased in frequency through the founding bottleneck effect in the Finnish population; occurring 19 times more frequently in Finns compared to non-Finnish Europeans and extremely rare in non-European populations (Table 1). These SNP alleles are in complete disequilibrium (*D*′ = 1.0) but poorly correlated (*r*^2^ = 0.037) due to their disparate frequencies. After conditioning on rs570516915, the rs7546104 variant showed residual evidence of association exceeding genome-wide significance (OR_cond_ = 1.75, *P*_cond_ = 3.98 × 10^−10^), supporting earlier evidence that rs7546104 tags an independently acting association signal^8^ (Supplementary Fig. 5c-d). Because of the high *D*′ value between the two alleles, reciprocal conditioning was not possible. However, the role of rs570516915 in CP susceptibility was already evident by its robust association with large effect size and location in an essential craniofacial gene enhancer. Further analysis of genotype effects on CP risk indicated that the frequency of CP cases rose significantly when the rs570516915 risk allele dosage on the ancestral risk haplotype tagged by rs7546104 was increased to the heterozygous or homozygous state (almost 10-fold or 4-fold, respectively; Supplementary Table 7). Thus, rs570516915 defines an independent CP risk haplotype that contributes greater, and stronger, risk specifically in the Finnish population.

Furthermore, the risk allele (G) of rs570516915 is tightly linked (*D*′ = 1.0, *r*^2^ = 0.0034) to the non-risk allele (also G) of the previously described common CL/P risk variant rs642961 in the same enhancer,^22^ suggesting that the association signals are independent of one another. Supporting this conclusion, the rs570516915 variant shows a significant association with CP alone, while rs642961 is strongly associated with CL and modestly associated with CLP.^22^ The risk allele A of rs642961, which has a frequency of 22% in the Finnish population, did not reach statistical significance in the Finnish cohort of CL/P cases evaluated here (*P* = 0.4), possibly because of the small sample size, and showed nonsignificant negative association with CP (OR = 0.71, 95% CI 0.57–0.89, *P* = 0.003).

### Replication in independent Finnish and Estonian cohorts

To replicate the association signals identified in the CP cohort, we performed GWAS in an independent sample of 165 CP cases and 199,935 populations controls from the FinnGen study (Data Freezes 8 through 12), and 71 CP cases and 198,973 ethnicity-matched population controls from the Estonian Biobank. The Estonian population is geographically and linguistically relatively close to the Finnish population, and many genetic associations in the Finnish population have been replicated in the Estonian Biobank.^23^ Risk allele of the MCS-9.7 variant rs570516915 also showed genome-wide significant association in the Finnish replication cohort (*P* = 8.82 × 10^−28^) and suggestive evidence of association in the Estonian cohort (*P* = 1.25 × 10^−5^) with the direction of effect consistent across all three cohorts. In a meta-analysis of the discovery and two replication datasets, the rs570516915 variant yielded a *P* value of 4.34 × 10^−64^. Summary statistics of the other two independent signals in the replication cohorts and meta-analysis results are shown in Table 1. Variants reaching *P* values less than 1.0 × 10^−4^ in these additional analyses are shown in Supplementary Tables 8, 9 and 10. Notably, in a small cohort of Estonian CL/P cases (n = 53), none of these variants showed significant association. In summary, the novel association of rs570516915 with CP was confirmed in independent datasets.

### Correlation of regional rs570516915 allele frequency with regional CP prevalence

To test whether the distribution of the risk allele tracked with CP prevalence across the 19 administrative regions of Finland, we estimated the distribution of allele frequency using region-level birthplace registry data of 306,678 FinnGen study participants (∼5.5% of Finland’s total population). We observed that the risk allele shows higher frequency in the eastern and northern regions of the country, reaching up to 2.7% (Fig. 3a and Supplementary Table 11), where the incidence of CP is also known to be higher compared to the western and southern regions of the country.^5–7^ The risk allele frequency in western and southern regions is lower than the average allele frequency in the entire country (i.e., 1.2%). We next estimated the prevalence of CP in each region of Finland using data from a large, nationwide registry study (FinRegistry, https://www.finregistry.fi). In electronic health records of 5,216,731 individuals, we identified 5,162 individuals with ICD codes related to non-syndromic CP. Strikingly, the CP prevalence also showed a geographic distribution that increased from south to north and from west to east (Fig. 3a and Supplementary Table 11). Most importantly, we found a strong and statistically significant correlation (Pearson’s *r* = 0.68, *P* < 0.001) between the regional distribution of the risk allele and CP prevalence (Fig. 3b). On the other hand, the frequency of the risk allele for the other two associated SNPs, rs41268753 (p.Thr454Met in *GRHL3*) and rs7546104 (*IRF6* locus), did not vary similarly across the 19 regions in Finland (data not shown). Therefore, of these three CP-associated SNPs, only rs570516915 appears to contribute to the regional distribution pattern of CP in Finland. This observation also supports our earlier conclusion that the effect of rs570516915 is independent of rs7546104.

**Fig. 3:**
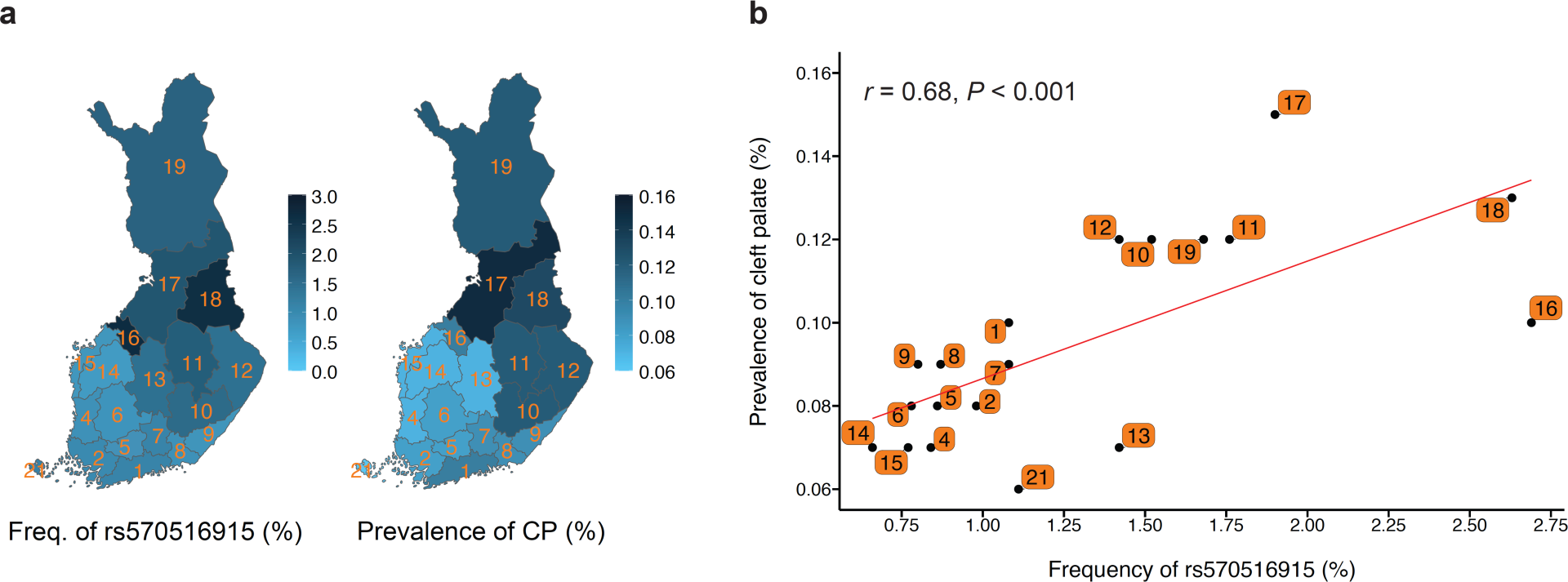
Geographic distribution and correlation of the rs570516915 variant allele frequency with regional prevalence of non-syndromic CP in Finland. **a**, Distribution of the rs570516915 G allele frequency and CP prevalence in 19 administrative regions of Finland. The variation in allele frequency and CP prevalence across distinct regions is illustrated by the intensity of the blue shading. Regions are labeled by their corresponding codes from Statistics Finland and shown in Supplementary Table 9. **b**, Correlation between the regional allele frequency of rs570516915 and prevalence of CP. Allele frequency was estimated using birthplace data of 306,678 FinnGen study participants. Regional CP prevalences were estimated using nationwide data on the first recorded addresses and CP related diagnosis codes of 5,216,731 individuals (5,162 with CP). Red line represents the ordinary least squares regression line. Pearson’s *r* and *P* value (two-sided) shown at the top of the plot indicates the strength and statistical significance of correlation. See Supplementary Table 11 for raw numbers.

We also estimated the overall impact of these three variants on CP in the Finnish population by calculating their individual and joint population attributable risks (PAR). Even though rs570516915 has a much lower frequency than rs41268753 and rs7546104 in Finland (4 times and 20 times lower, respectively, Table 1), its PAR (16.2%) is comparable to that of rs41268753 (14.4%) and is nearly half of rs7546104 (27.3%). The latter was calculated using an adjusted odds ratio from the conditional analysis and frequency of the risk allele (22.9%) that is not linked to rs570516915. The reason for the strong PAR by rs570516915 is its large effect size. The large PAR for rs7546104 is likely overestimated since the frequency, effect size and the number of the true causal variant(s) tagged by rs7546104 are unknown. The combined PAR of these three variants on CP is 47.8%.

### rs570516915 disrupts regulatory activity of the *IRF6* enhancer MCS-9.7

Six SNPs were in strong LD (*r*^2^ > 0.8) with the lead SNP rs570516915. To prioritize these SNPs for possible regulatory variants, we examined their position relative to chromatin marks shown to correlate with active enhancer function including: i) ATAC-seq peaks, which reveal open chromatin, from the HIOEC oral epithelial cells and the HEPM oral mesenchymal cells,^41^ ii) H3K27Ac peaks, revealing active enhancers and promoters, from the HIOEC cells,^41^ and iii) aggregate chromatin modification marks from 9 principal cell types from the ENCODE project^39^ and from human embryonic facial enhancers datasets.^40^ Of these seven SNPs, the lead SNP and one other fall into regions with chromatin marks consistent with enhancer activity in epithelial cells (HIOEC and NHEK) (Fig. 4a and Supplementary Fig. 7c). As mentioned above, the lead SNP rs570516915 is located in MCS-9.7, a known enhancer for *IRF6*^22^ in which three variants have been associated with OFCs previously.^8,22,52^ The second SNP, rs556188853, is located 58 kb downstream from *IRF6*. We next engineered 701 bp DNA fragments including the ATAC-seq peak in HIOEC cells containing rs570516915 or rs556188853 and harboring either the risk-associated or non-risk associated allele of the SNP into a luciferase-based reporter vector (Supplementary Notes, Supplementary Fig. 8). We independently transfected these constructs into primary human neonatal epidermal keratinocytes (HEKn). The vector harboring MCS-9.7 with the ancestral, non-risk allele of rs570516915 drove luciferase levels above background. However, the vector harboring the derived risk allele of rs570516915 showed significantly reduced activity relative to the one harboring the non-risk allele (*P* = 0.0002) (Fig. 4b). By contrast, the vectors harboring the region surrounding rs556188853 did not drive luciferase levels above background, and there was no significant difference in luciferase levels between the vectors harboring the two alleles (Fig. 4b). These results support that the risk allele at rs570516915 is functional by reducing the activity of the MCS-9.7 enhancer in oral keratinocytes.

**Fig. 4:**
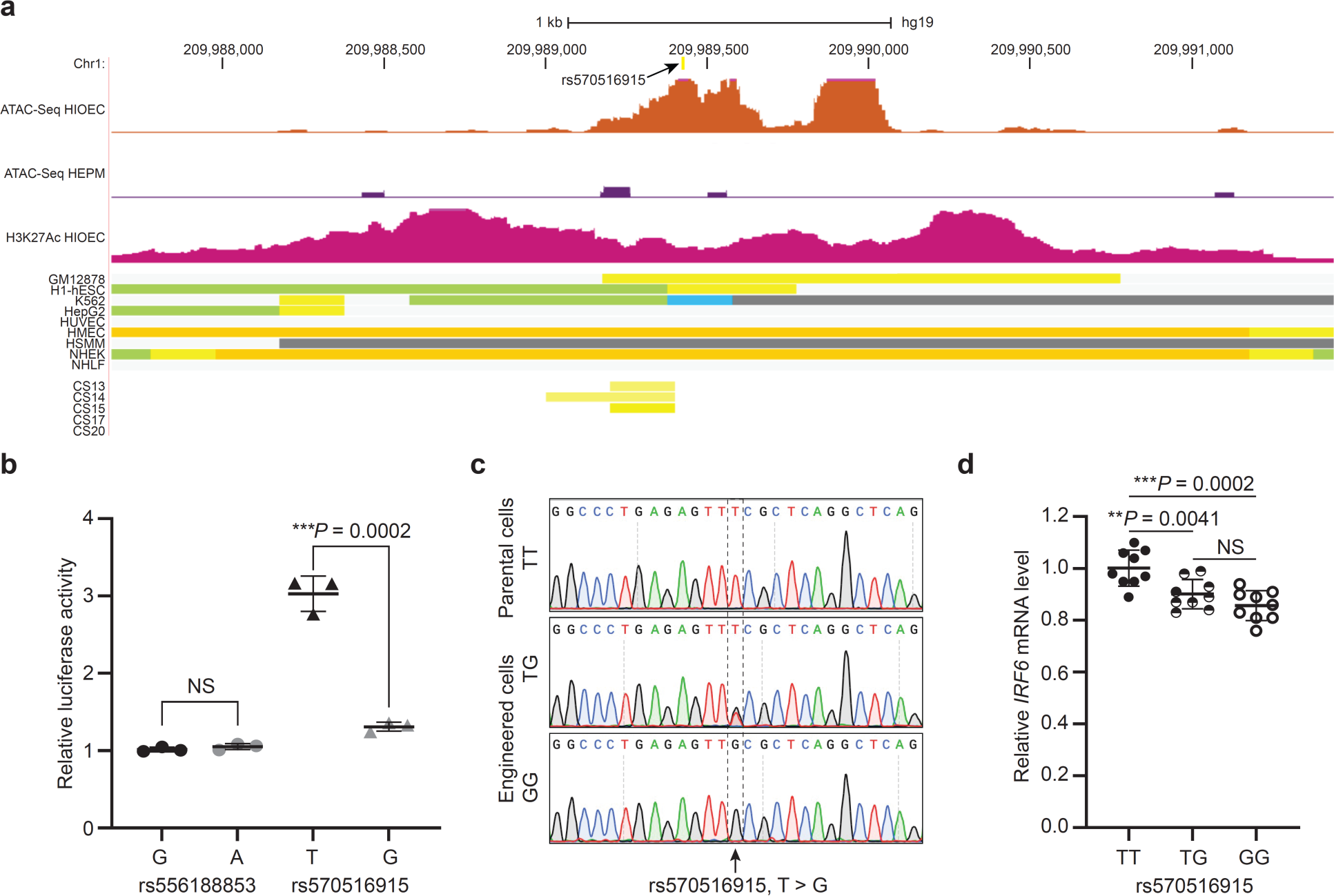
The rs570516915 variant disrupts enhancer activity of MCS-9.7. **a**, Browser view of the human genome, GRCh37/hg19, focused on the rs570516915 variant. Top two tracks represent open chromatin peaks detected by ATAC-Seq in HIOEC and HEPM cells, respectively. Next track shows H3K27Ac marks illustrating rs570516915 as a part of enhancer element. Following color coded bars represent chromatin status revealed by ChIP-Seq to various chromatin marks from the ENCODE Project cell lines and facial explants from human embryos at Carnegie stage (CS) 13-20, encompassing the time when palate shelves fuse, where orange and yellow bars represent the active and weak enhancer element, respectively; blue, insulator; light green, weak transcribed; grey, Polycomb repressed; light grey, heterochromatin/repetitive; GM12878, B-cell derived cell line; ESC, embryonic stem cells; K562, myelogenous leukemia; HepG2, liver cancer; HUVEC, human umbilical vein endothelial cells; HMEC, human mammary epithelial cells; HSMM, human skeletal muscle myoblasts; NHEK, normal human epidermal keratinocytes; NHLF, normal human lung fibroblasts, CS13-CS20 are facial explants from human embryos. **b**, Scattered dot plot of relative luciferase activity for non-risk and risk alleles of rs556188853 and rs570516915 in HEKn cells. Data are represented as mean values +/− s. d. from three independent experiments. Statistical significance is determined by Student’s *t*-test. *P* value (two-tailed) is indicated on the plot and NS represents non-significant (*P* = 0.2452). **c**, Chromatograms illustrating the three genotypes (TT, TG and GG) of rs570516915 in iPSCs generated by CRISPR-Cas9 mediated homology-directed repair. **d**, Scattered dot plot of relative levels of *IRF6* mRNA in edited vs parental iOECs assessed by qRT-PCR. Expression levels of *IRF6* are normalized against *ACTB, GAPDH, HPRT, UBC* and *CDH1*. Data are represented as mean values +/− s. d. from nine replicates of cells harboring each genotype, as indicated in the plot. Statistical significance is determined by Student’s *t*-test (two-tailed). NS represents non-significant (*P* = 0.1212).

To test the *in vivo* effect of the risk allele on enhancer activity, we performed a transgenic F0 embryo assay with the MCS-9.7 enhancer fused to the *lacZ* reporter gene^45^ (Supplementary Notes). We produced embryos in three experimental batches and in each batch both single and tandem insertions were obtained, providing six sets of embryos with variable numbers of embryos (Supplementary Fig. 1). At E13.5, most embryos carrying the MCS-9.7*-lacZ* transgene with either the risk and the non-risk allele showed staining in the ectoderm of face, torso, limbs, eye and pinna, consistent with previous staining patterns.^22,53^ X-gal staining, which showed variation both between and within batches, was assessed by blinded independent observers. In three of the sets (1S, 1T, 2S), non-risk allele carrying embryos were scored on average as having higher X-gal staining intensity than risk-allele carrying embryos (Supplementary Fig. 1 and Supplementary Table 1). However, in the other three sets (2T, 3S, 3T) the results were less distinct, and this technical limitation should be taken into account when interpreting the data. Nonetheless, a Wilcoxon Signed Ranks test of the rankings from all six sets indicated that the non-risk allele carrying embryos were ranked significantly higher compared to the risk allele carrying embryos (*P* = 0.027), providing support for an *in vivo* effect of the risk allele on MCS-9.7 activity.

We next determined the effect of rs570516915 in its native chromatin context on the expression of *IRF6* in embryonic oral epithelial cells derived from induced pluripotent stem cells (iPSCs) (i.e., induced oral epithelial cells, iOECs). Sequencing showed that WTC11 iPSCs were homozygous for the non-risk allele (i.e., TT) of rs570516915. We transfected these cells with appropriate CRISPR-Cas9 reagents to render them heterozygous (i.e., GT) or homozygous (i.e., GG) for the risk allele and isolated three independent clones of both genotypes (Fig. 4c and Supplementary Figs. 2 and 3). We subjected iPSCs of each genotype to a differentiation protocol that converts them to embryonic oral epithelium^46^ (Supplementary Fig. 3). Quantitative RT-PCR and immunostaining revealed induction of multiple epithelial (*KRT8*, *KRT18*, *CDH1*, *IRF6* and *TP63*) and oral markers (*PITX1* and *PITX2*) in iOECs relative to in undifferentiated parental iPSCs (Supplementary Figs. 9 and 10). Interestingly, *IRF6* mRNA levels were lower in cells heterozygous or homozygous for the risk allele compared with the cells homozygous for the non-risk allele (Fig. 4d). These findings provide evidence that the risk allele at rs570516915 directly affects risk for CP by reducing steady state levels of *IRF6* mRNA.

### rs570516915 disrupts the autoregulatory activity of the *IRF6* gene

Motif analysis of the conserved sequences surrounding rs570516915 revealed that the risk-associated allele disrupts a consensus binding site for IRF6, a transcription factor known to bind DNA (Fig. 5a and Supplementary Fig. 11). Prior *in vitro*^54^ and *in vivo*^55^ studies showed that IRF6 regulates its own expression, suggesting that the risk-associated allele of rs570516915 disrupts IRF6 autoregulation. To test this hypothesis, we performed chromatin immunoprecipitation with anti-H3K27Ac and with anti-IRF6 antibodies followed by PCR with primers flanking rs570516915 (ChIP-qPCR), or with primers at a control locus lacking evidence of enhancer activity in epithelial cells, in iOECs that were heterozygous for the risk and non-risk alleles of rs570516915. Quantitative PCR revealed much higher binding of H3K27Ac and of IRF6 at MCS-9.7 versus at the control locus in iOECs (Fig. 5b and 5c). Interestingly, sequencing of the amplified DNA precipitated by anti-H3K27Ac and by anti-IRF6 revealed that both antibodies preferentially precipitated the chromosome harboring the non-risk allele (Fig. 5d); the same observation was made in two other biological replicates (Supplementary Fig. 12). These data are consistent with a mechanism whereby rs570516915 disrupts the binding of IRF6 protein at a critical site in MCS-9.7 to destabilize a positive feedback loop for *IRF6* expression that is required for proper palatal development (Supplementary Fig. 13).

**Fig. 5:**
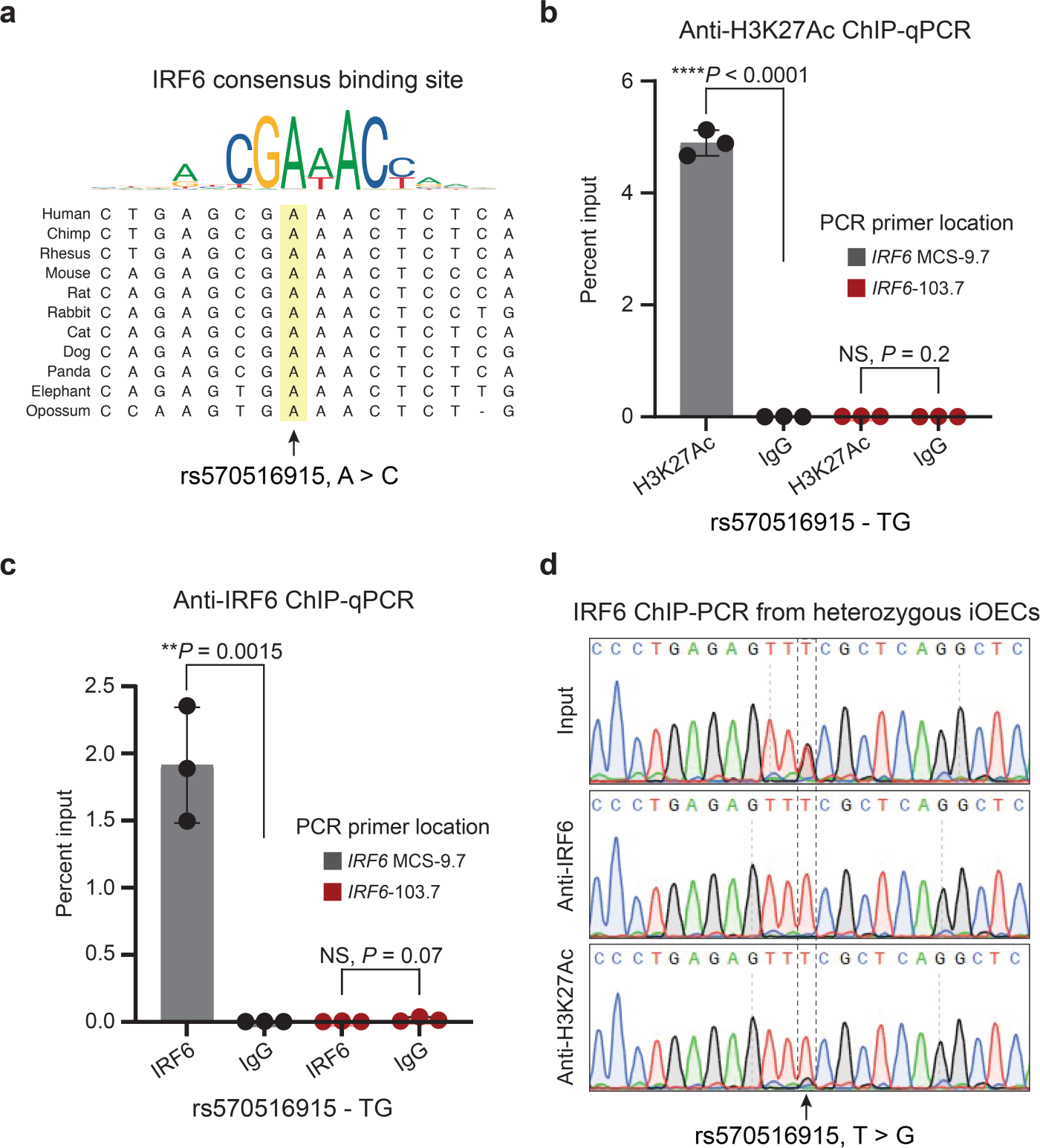
rs570516915 disrupts an IRF6 binding site and perturbs positive autoregulation of *IRF6* expression via the MCS-9.7 enhancer. **a**, Consensus IRF6 binding motif from the JASPAR database of transcription factor DNA-binding preferences (Matrix ID: PB0036.1) and alignment of the variant site in different species, which shows that the ancestral allele A is completely conserved in mammals and is critical for binding of IRF6. Note that this A corresponds to the T allele of rs570516915 on the opposite DNA strand. **b** and **c**, Percent input identified by ChIP-qPCR for anti-H3K27Ac and anti-IRF6 respectively in iOECs heterozygous for rs570516915 using primers specific to the MCS-9.7 enhancer site or, as a negative control, to a region 103.7 kb upstream *IRF6* transcription start site that did not harbor active elements identified from ATAC-Seq and H3K27Ac ChIP-Seq in HIOEC or NHEK cells and devoid of predicted IRF6 binding sites. Error bars refer to 3 ChIP replicates and expressed as mean values +/− s. d. Statistical significance is determined by Student’s *t*-test. *P* value (two-tailed) is indicated on the plot and NS represents non-significant. **d**, Sequencing of anti-IRF6 and anti-H3K27Ac ChIP-PCR product of cells heterozygous for rs570516915.

## Discussion

Following multiple waves of settlement after the last glacial period, several genetic founder and bottleneck events have occurred in Finland. Despite the considerable expansion of the population since the early 1800s, regions of genetic isolation have persisted.^56,57^ Consequently due to strong genetic drift, individuals of Finnish ancestry have numerous deleterious alleles of clinical significance at a relatively high frequency compared to non-Finnish Europeans.^58^ This is well-exemplified by a group of rare monogenic diseases, also known as the “Finnish disease heritage”, that exists at a higher frequency in Finland than in other Nordic countries,^59,60^ which all share similar living standards, climate, infectious diseases, diet, and cultural habits. Examples of strong regional concentration of complex diseases also exist; for instance, schizophrenia, cognitive impairment, and familial hypercholesterolemia are more prevalent in the northeastern region of the Finland than in other regions and show association with regionally enriched genetic variation.^61,62^ It was postulated before the genomics era that the relatively high incidence of CP in Finland compared to other Nordic countries, which all have similar total incidence of clefts as Finland, is likely to result from the distinct components of genetic heritage of Finns.^4^ Here we have shown the association of three independent SNPs with high allele frequencies in Finland compared to other populations which explains the high incidence rate of CP in Finland.

Further, we found evidence that one of the three SNPs, rs570516915, directly elevates risk for CP by disrupting positive regulation by IRF6 of its own expression. Previously, chromatin immunoprecipitation combined with next generation sequencing (ChIP-seq) in primary human keratinocytes identified a 245 bp sequence within the MCS-9.7 enhancer of *IRF6* where the IRF6 protein binds.^54^ Furthermore, an *in vivo* study showed that Irf6 is required for enhancer activity of MCS-9.7 in tissues derived from ectodermal germ layer during lip and palate development in mouse embryos.^55^ Here, we identified an OFC-associated risk variant in a conserved IRF6 binding site within MCS-9.7. The risk allele of this SNP significantly affected MCS-9.7 enhancer activity in transient luciferase reporter assays in keratinocytes *in vitro.* As a limitation, we did not see a fully consistent effect in a transient transgenic mouse reporter assay, in contrast to a rare VWS-associated variant in MCS-9.7, which had a strong effect in this assay.^52^ The differential effects in this assay likely reflect the small and large effects that these variants have on OFC risk. Nonetheless, altering rs570516915 within the genome of induced oral epithelial cells, where basal *IRF6* expression levels are higher, was sufficient to affect binding of IRF6 and expression of *IRF6*.

These and the published data mentioned above are consistent with a model where autoregulation of *IRF6* occurs through direct binding of the IRF6 protein to the conserved IRF6 binding site in MCS-9.7, and that the risk allele of rs570516915 disrupts this autoregulation. However, we cannot exclude more complex scenarios. For example, we note that while the conserved IRF6 binding site that contains rs570516915 is located within MCS-9.7, it is located 40 bp outside of the previously described 245 bp region of chromatin bound by IRF6.^54^ Similarly, it is located 22 bp from a region of chromatin marked as active enhancer in human embryonic faces collected at Carnegie stages 13-15 (Supplementary Fig. 14).^40^ Thus, indirect mechanisms of autoregulation cannot be excluded. *IRF6* stands out as one of two genes connected to both syndromic and non-syndromic forms of CL/P and CP, and remarkably, different variants within the MCS-9.7 enhancer of *IRF6* are associated with this phenotypic spectrum. To date, rare pathogenic mutations in multiple genes are known to cause syndromic forms of CL/P and CP (81 and 78 genes, respectively),^63^ and common alleles at multiple loci increase risk for non-syndromic CL/P ^8–11,64–72^ and CP^8–10,12,13^ (47, and 15 respectively). Rare loss-of-function variants in *IRF6* cause Van der Woude syndrome,^73^ where individuals with CL/P and individuals with CP can be found in the same family. Such “mixed cleft” families are exceptional, as development of the lip and palate are embryologically and generally, epidemiologically distinct.^74–76^ In addition, this and prior studies have identified common variants at the *IRF6* locus that are associated with non-syndromic CL/P^22,77^ or with CP.^8,9,11^ A single nucleotide duplication, chr1:g.209816135dup (GRCh38/hg38), aka 350dupA^52^, was found in one VWS family, where two members had CL/P and one member had CP. A common SNP, rs642961 (chr1:209815925), was shown to be associated with non-syndromic CL/P,^22^ and now the low-frequency Finnish-enriched SNP rs570516915 (chr1:209816078) shows strong association with non-syndromic CP. All three variants are located within a 212 bp segment of the MCS-9.7 enhancer (Supplementary Fig. 14). In addition, a GWAS in the Han Chinese population identified rs72741048 (chr1:209815747) as a lead SNP associated both with CL and CP, 331 bp upstream of rs570516915, but the pathogenic variant remains to be identified.^8^

How do different mutations in the same enhancer predispose to CP versus to CL/P? In the palate, MCS-9.7 is active in basal and superficial (periderm) epithelial layers, replicating endogenous *Irf6* expression patterns.^53^ Both layers are important during fusion of the palate shelves ^21,78^ and during fusion of the lip at the lambdoidal junction.^79^ Nonetheless, it is possible that different mutations in MCS-9.7 differentially affect its enhancer activity in basal and periderm layers and thereby differentially affect risk for CP versus CL/P. In this light, we favor periderm as the relevant tissue where rs570516915 contributes risk to CP. Our rationale is based on the overlapping genetics and function between *IRF6* and *GRHL3*. Specifically, as observed with the common risk variant^13^ in *GRHL3*, rs570516915 is associated with non-syndromic CP but not with non-syndromic CL/P. Also, while rare variants in both *IRF6* and *GRHL3* cause VWS, individuals with a mutation in *GRHL3* are more likely to have a CP, rather than CL/P.^17^ Finally, the primary pathologies in mice that lack either *Irf6*^18,19^ or *Grhl3*^17^ are abnormal intraoral epithelial adhesions and failure of periderm formation or function, which hinder development of the palatal shelves and can cause cleft palate.^78^ Precise genome editing at these conserved variant sites in mouse models may reveal the distinct effects of the variants on *Irf6* expression in periderm versus basal layers. In conclusion, the MCS-9.7 regulatory region may represent a mutational “hot” spot for both rare and common genetic variation that cause and contribute risk for the syndromic and non-syndromic forms of OFCs.

## Data availability

Summary results of variants with *P* values less than 1.0 × 10^−4^ are provided in Supplementary Tables 4, 5, 6, 8, 9 and 10. Full GWAS summary statistics from the FinnGen discovery cohort are available for download from https://www.finngen.fi/en/access_results (DF7 - June 1, 2022).

## FinnGen ethics statement

The Coordinating Ethics Committee of the Hospital District of Helsinki and Uusimaa (HUS) statement number for the FinnGen study is Nr HUS/990/2017. The FinnGen study is approved by the Finnish Institute for Health and Welfare (THL, permit numbers [PN]: THL/2031/6.02.00/2017, THL/1101/5.05.00/2017, THL/341/6.02.00/2018, THL/2222/6.02.00/2018, THL/283/6.02.00/2019, THL/1721/5.05.00/2019 and THL/1524/5.05.00/2020); the Digital and Population Data Services Agency (PN: VRK43431/2017-3, VRK/6909/2018-3, VRK/4415/2019-3); the Social Insurance Institution (PN: KELA 58/522/2017, KELA 131/522/2018, KELA 70/522/2019, KELA 98/522/2019, KELA 134/522/2019, KELA 138/522/2019, KELA 2/522/2020, KELA 16/522/2020); Findata (PN: THL/2364/14.02/2020, THL/4055/14.06.00/2020, THL/3433/14.06.00/2020, THL/4432/14.06/2020, THL/5189/14.06/2020, THL/5894/14.06.00/2020, THL/6619/14.06.00/2020, THL/209/14.06.00/2021, THL/688/14.06.00/2021, THL/1284/14.06.00/2021, THL/1965/14.06.00/2021, THL/5546/14.02.00/2020, THL/2658/14.06.00/2021, THL/4235/14.06.00/2021); Statistics Finland (PN: TK-53-1041-17 and TK/143/07.03.00/2020 [earlier TK-53-90-20], TK/1735/07.03.00/2021, TK/3112/07.03.00/2021) and the Finnish Registry for Kidney Diseases (permission/extract from the meeting minutes on July 4, 2019). The Biobank Access Decisions for FinnGen samples and data include: THL Biobank BB2017_55, BB2017_111, BB2018_19, BB_2018_34, BB_2018_67, BB2018_71, BB2019_7, BB2019_8, BB2019_26, BB2020_1, BB2021_65, Finnish Red Cross Blood Service Biobank 7.12.2017, Helsinki Biobank HUS/359/2017, HUS/248/2020, HUS/430/2021 §28, §29, HUS/150/2022 §12, §13, §14, §15, §16, §17, §18, §23, §58, §59, HUS/128/2023 §18, Auria Biobank AB17-5154 and amendment #1 (August 17, 2020) and amendments BB_2021-0140, BB_2021-0156 (August 26, 2021, February 2, 2022), BB_2021-0169, BB_2021-0179, BB_2021-0161, AB20-5926 and amendment #1 (April 23, 2020) and its modifications (September 22, 2021), BB_2022-0262, BB_2022-0256, Biobank Borealis of Northern Finland_2017_1013, 2021_5010, 2021_5010 Amendment, 2021_5018, 2021_5018 Amendment, 2021_5015, 2021_5015 Amendment, 2021_5015 Amendment_2, 2021_5023, 2021_5023 Amendment, 2021_5023 Amendment_2, 2021_5017, 2021_5017 Amendment, 2022_6001, 2022_6001 Amendment, 2022_6006 Amendment, 2022_6006 Amendment, 2022_6006 Amendment_2, BB22-0067, 2022_0262, 2022_0262 Amendment, Biobank of Eastern Finland 1186/2018 and amendment 22§/2020, 53§/2021, 13§/2022, 14§/2022, 15§/2022, 27§/2022, 28§/2022, 29§/2022, 33§/2022, 35§/2022, 36§/2022, 37§/2022, 39§/2022, 7§/2023, 32§/2023, 33§/2023, 34§/2023, 35§/2023, 36§/2023, 37§/2023, 38§/2023, 39§/2023, 40§/2023, 41§/2023, Finnish Clinical Biobank Tampere MH0004 and amendments (21.02.2020 & 06.10.2020), BB2021-0140 8§/2021, 9§/2021, §9/2022, §10/2022, §12/2022, 13§/2022, §20/2022, §21/2022, §22/2022, §23/2022, 28§/2022, 29§/2022, 30§/2022, 31§/2022, 32§/2022, 38§/2022, 40§/2022, 42§/2022, 1§/2023, Central Finland Biobank 1-2017, BB_2021-0161, BB_2021-0169, BB_2021-0179, BB_2021-0170, BB_2022-0256, BB_2022-0262, BB22-0067, decision allowing to continue data processing until August 31, 2024 for projects: BB_2021-0179, BB22-0067,BB_2022-0262, BB_2021-0170, BB_2021-0164, BB_2021-0161, and BB_2021-0169, and Terveystalo Biobank STB 2018001 and amendment August 25, 2020, Finnish Hematological Registry and Clinical Biobank decision June 18, 2021, Arctic biobank P0844: ARC_2021_1001.

## Acknowledgements

The FinnGen project is funded by two grants from Business Finland (HUS 4685/31/2016 and UH 4386/31/2016) and the following industry partners: AbbVie Inc., AstraZeneca UK Ltd, Biogen MA Inc., Bristol Myers Squibb (and Celgene Corporation & Celgene International II Sàrl), Genentech Inc., Merck Sharp & Dohme LCC, Pfizer Inc., GlaxoSmithKline Intellectual Property Development Ltd., Sanofi US Services Inc., Maze Therapeutics Inc., Janssen Biotech Inc, Novartis Pharma AG, and Boehringer Ingelheim International GmbH. Following biobanks are acknowledged for delivering biobank samples to FinnGen: Auria Biobank, THL Biobank, Helsinki Biobank, Biobank Borealis of Northern Finland, Finnish Clinical Biobank Tampere, Biobank of Eastern Finland, Central Finland Biobank, Finnish Red Cross Blood Service Biobank, Terveystalo Biobank and Arctic Biobank. All Finnish Biobanks are members of BBMRI.fi infrastructure. Finnish Biobank Cooperative – FINBB, is the coordinator of BBMRI-ERIC operations in Finland. The Finnish biobank data can be accessed through the Fingenious^®^ services managed by FINBB. FinRegistry is a collaboration project of the THL and the Data Science Genetic Epidemiology research group at the Institute for Molecular Medicine Finland (FIMM), University of Helsinki. The FinRegistry project has received the following approvals for data access from THL (THL/1776/6.02.00/2019 and subsequent amendments), DVV (VRK/5722/2019-2), Finnish Center for Pension (ETK/SUTI 22003) and Statistics Finland (TK-53-1451-19). The FinRegistry project has received IRB approval from THL (Kokous 7/2019). The FinRegistry project has received funding from the European Research Council (ERC) under the European Union’s Horizon 2020 research and innovation program (grant agreement No 945733), starting grant AI-Prevent. This project was also partially funded by grants from the U.S. National Institutes of Health, DE027362 (R.A.C.), DE023575 (R.A.C. and B.C.S.), HG010855 (KB) and CA246503 (KB). We would like to acknowledge support from the Centre of Excellence for Genomics and Translational Medicine (SLTMR16142T/TK142), and the Erasmus+ program of the European Union (2018-1-EL01-KA202-047907; 2018-1-SE01-KA202-039066). W.D.F. acknowledges support from the NIGMS grant R15-GM122030-01. We thank Dr. Akira Kinoshita, Nagasaki University, for providing the anti-IRF6 antibody. Finally, we would like to thank all participants and investigators of the FinnGen study and the Estonian Biobank.

