## Supplementary Material for "High incidence and geographic distribution of cleft palate cases in Finland are associated with a regulatory variant in *IRF6*"

**Supplementary Figures**

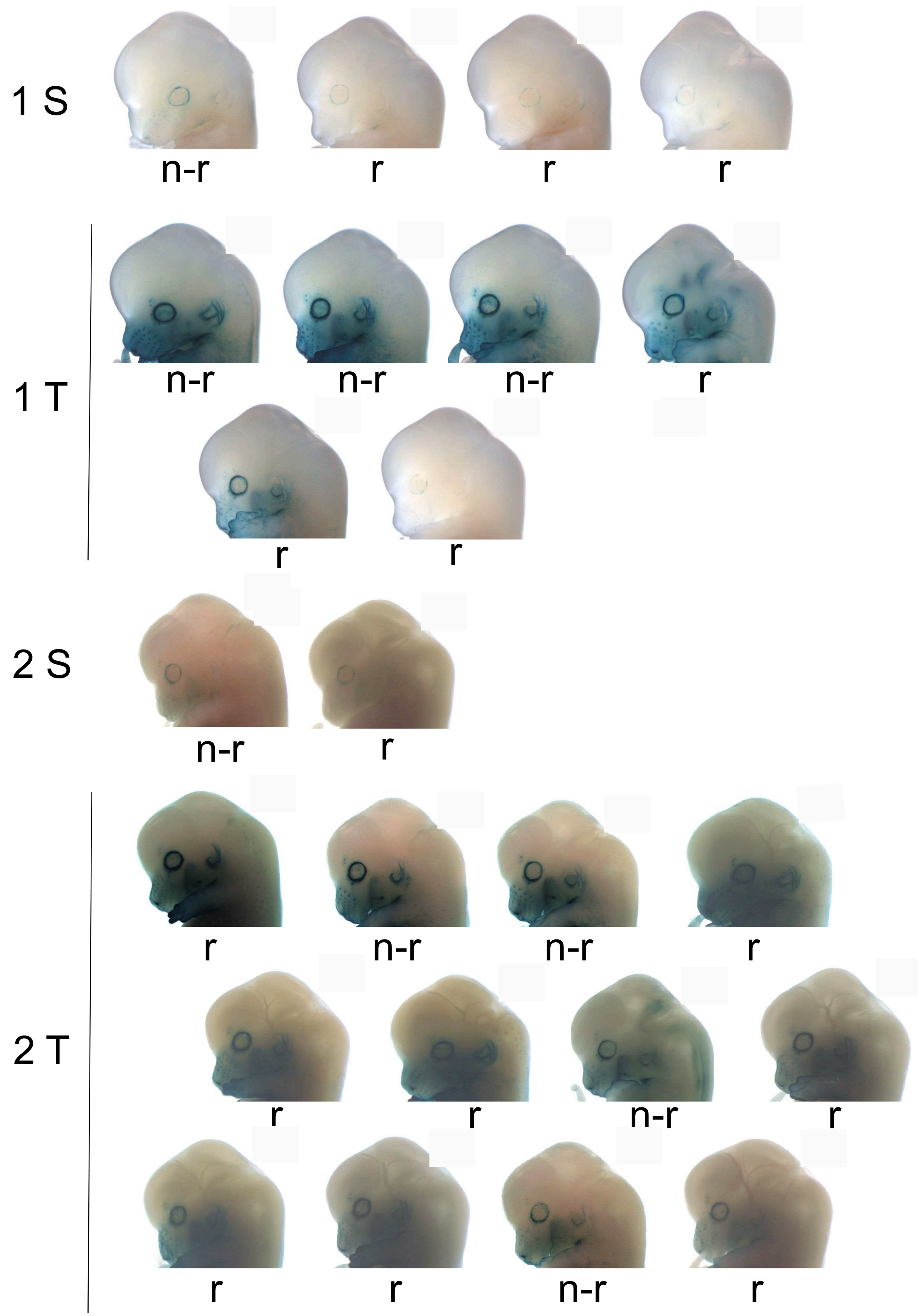

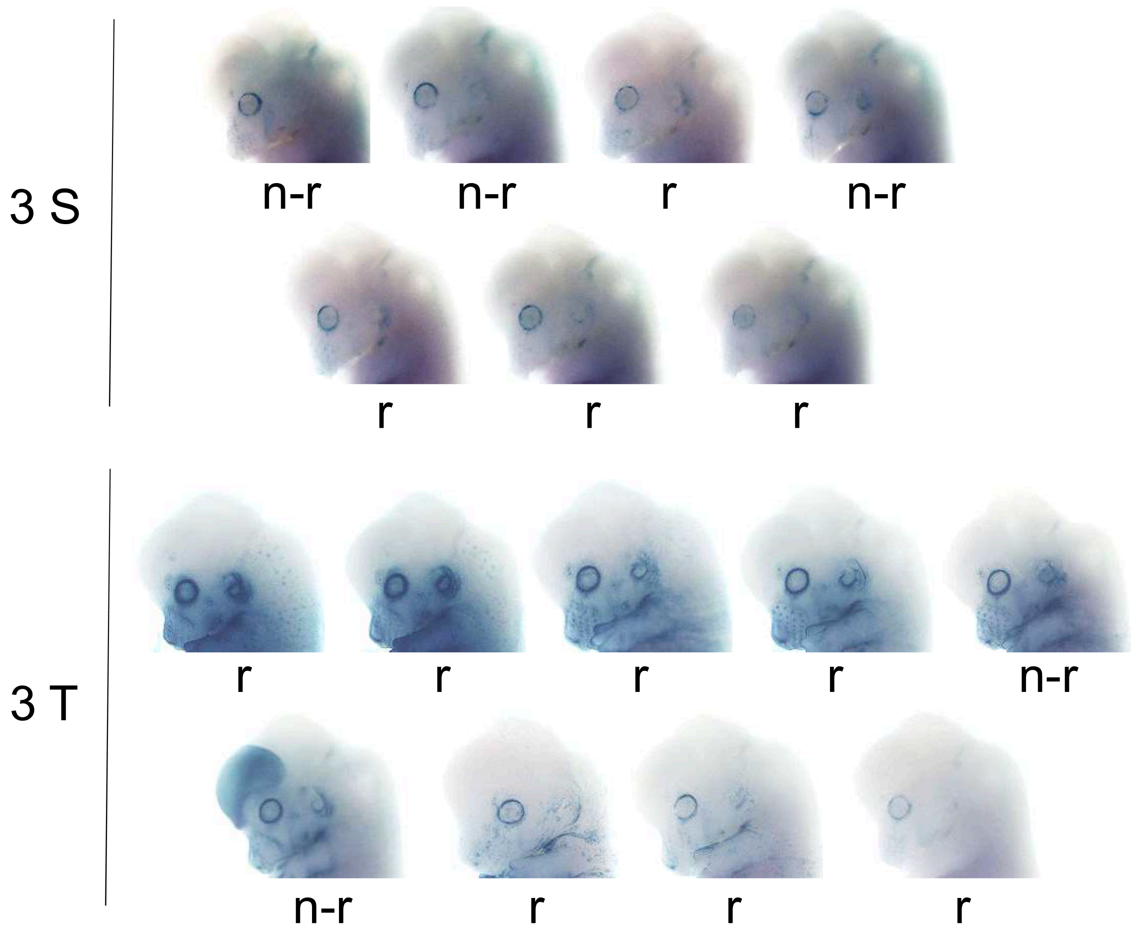


**Supplementary Fig. 1: Heads of X-gal-stained transgenic reporter embryos at E13.5 showing effect on staining of inserted non-risk and risk alleles of rs570516915.** Embryos were divided into six sets which consisted of single (S) or tandem (T) insertions from three experimental batches (1, 2 and 3). The heads are ordered according to consensuses from blinded assessments from strongest to weakest expression. Images of heads of sets 3S and 3T were adjusted similarly in each set for brightness and contrast. n-r, non-risk; r, risk.

Guide RNA and repair template sequences

**gRNA**

**Genome with T allele**

AGGCCCTGAGAGTTTCGCTC

XXXXXXTGGAGCTTTGGGGCCTGGGAACCTCTCTACCTGCGTCAATGTCTGGAGGCCCTGAGAGTTTCGCTCAGGCTCAGAGCAGGCATCGCAACCTCCCAGTTACTATTCTGTGCTGTGGCAAGXXXXXX

PAM

**HDR template with G allele**

TGGAGCTTTGGGGCCTGGGAACCTCTCTACCTGCGTCAATGTCTGGAGGCCCTGAGAGTTGCGCTCAGGCTCAGAGCAGGCATCGCAACCTCCCAGTTACTATTCTGTGCTGTGGCAAG

**Supplementary Fig. 2: Sequences of the guide RNA (gRNA) and homology-directed repair (HDR) template DNA used for CRISPR-Cas9 editing of the rs570516915 variant sequence.** PAM represents the protospacer adjacent motif for the guide used in the study.

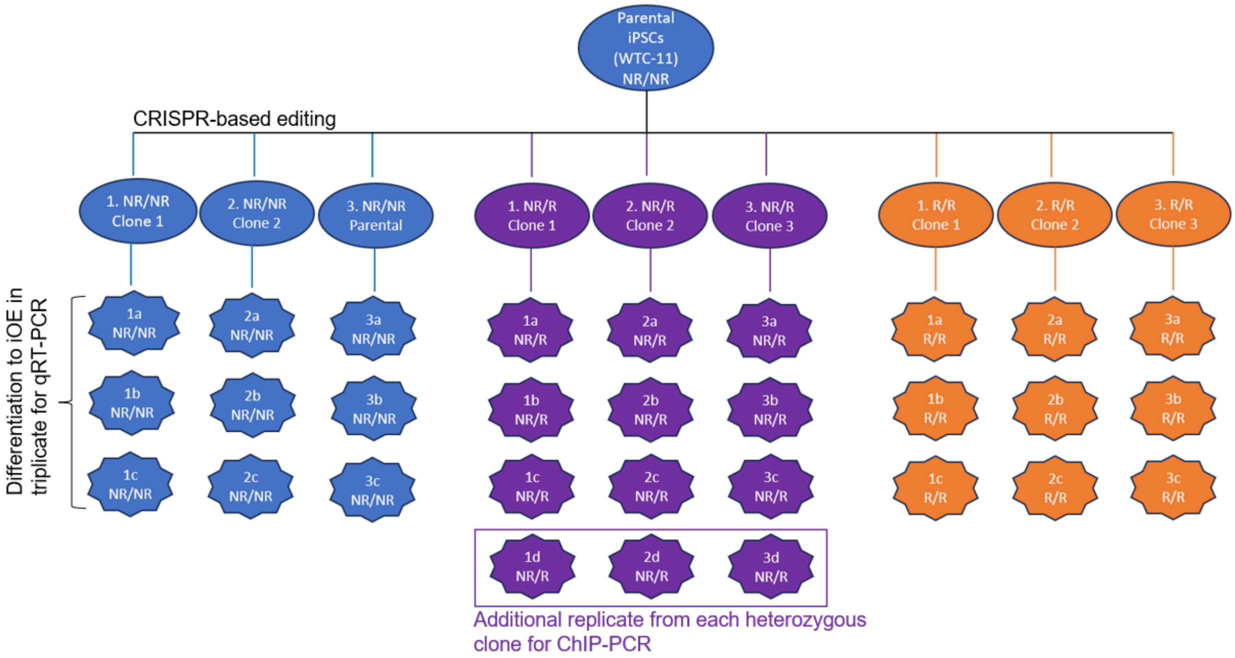


**Supplementary Fig. 3: Strategy for *in vitro* cell culture experiments:** Parental induced pluripotent stem cells (iPSCs) WTC-11, homozygous for non-risk allele of rs570516915 (TT-NR/NR), were edited to be heterozygous (TG-NR/R) or homozygous for the risk allele (GG-R/R) and subjected to a 10-day differentiation protocol to generate induced oral epithelial cells (iOECs). NR, non-risk allele; R, risk allele.

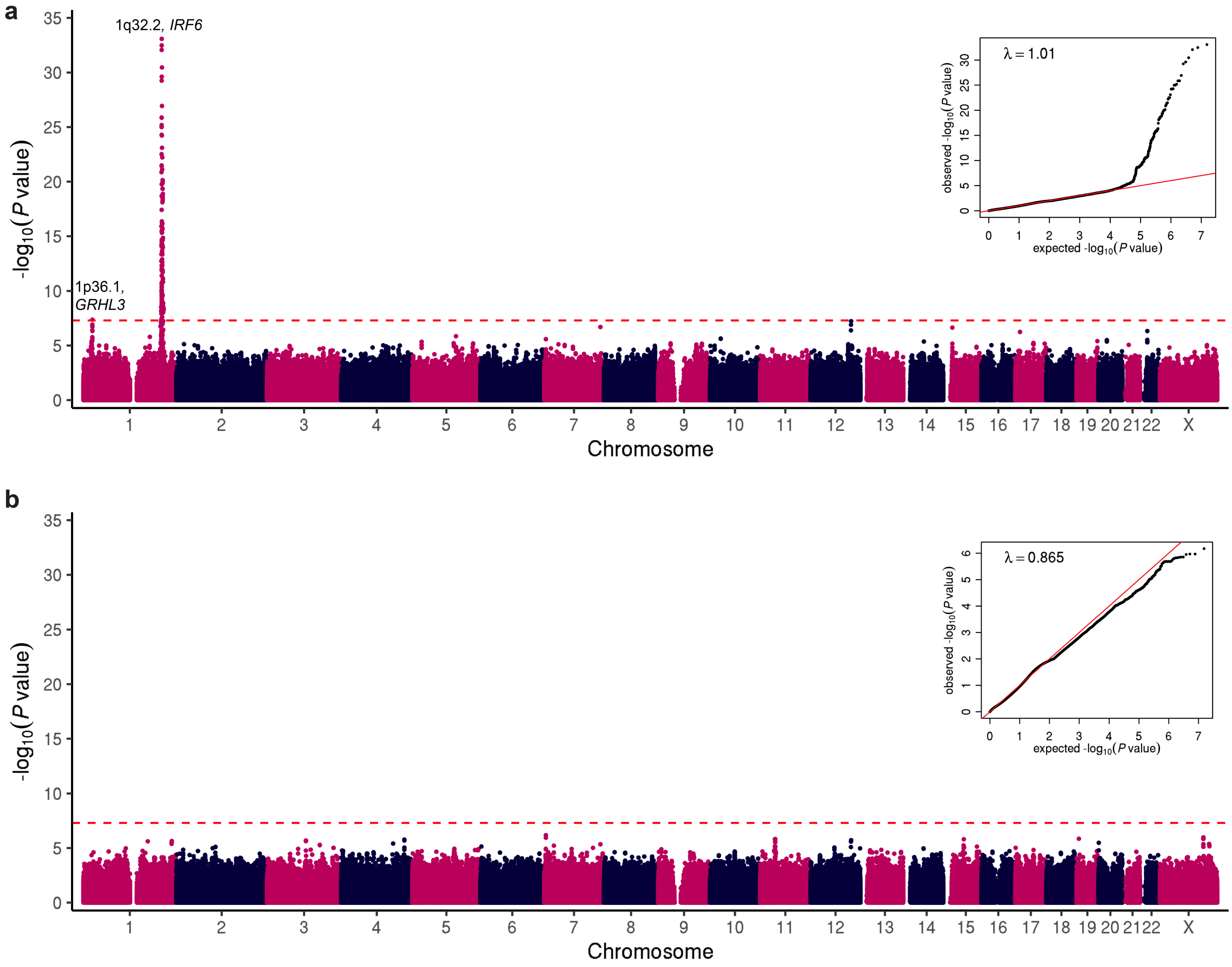


**Supplementary Fig. 4: Manhattan plots showing GWAS results of: a**, 355 cases with non-syndromic OFC (CL, CLP, and CP combined) and 308,799 population controls and, **b**, 151 cases with CL and CLP combined and 308,799 population controls. Negative $\log_{10}$ *P* values are plotted for each variant against their chromosomal coordinates provided in the human genome build GRCh38/hg38. Two-sided *P* values were estimated using Firth regression and not adjusted for multiple hypothesis testing. Red dashed line represents the threshold for genome-wide significance ($P = 5 \times 10^{-8}$) after Bonferroni correction for multiple testing. A Q-Q plot is shown in the inset panel, where the observed *P* values are plotted against the expected *P* values under null distribution (red line).

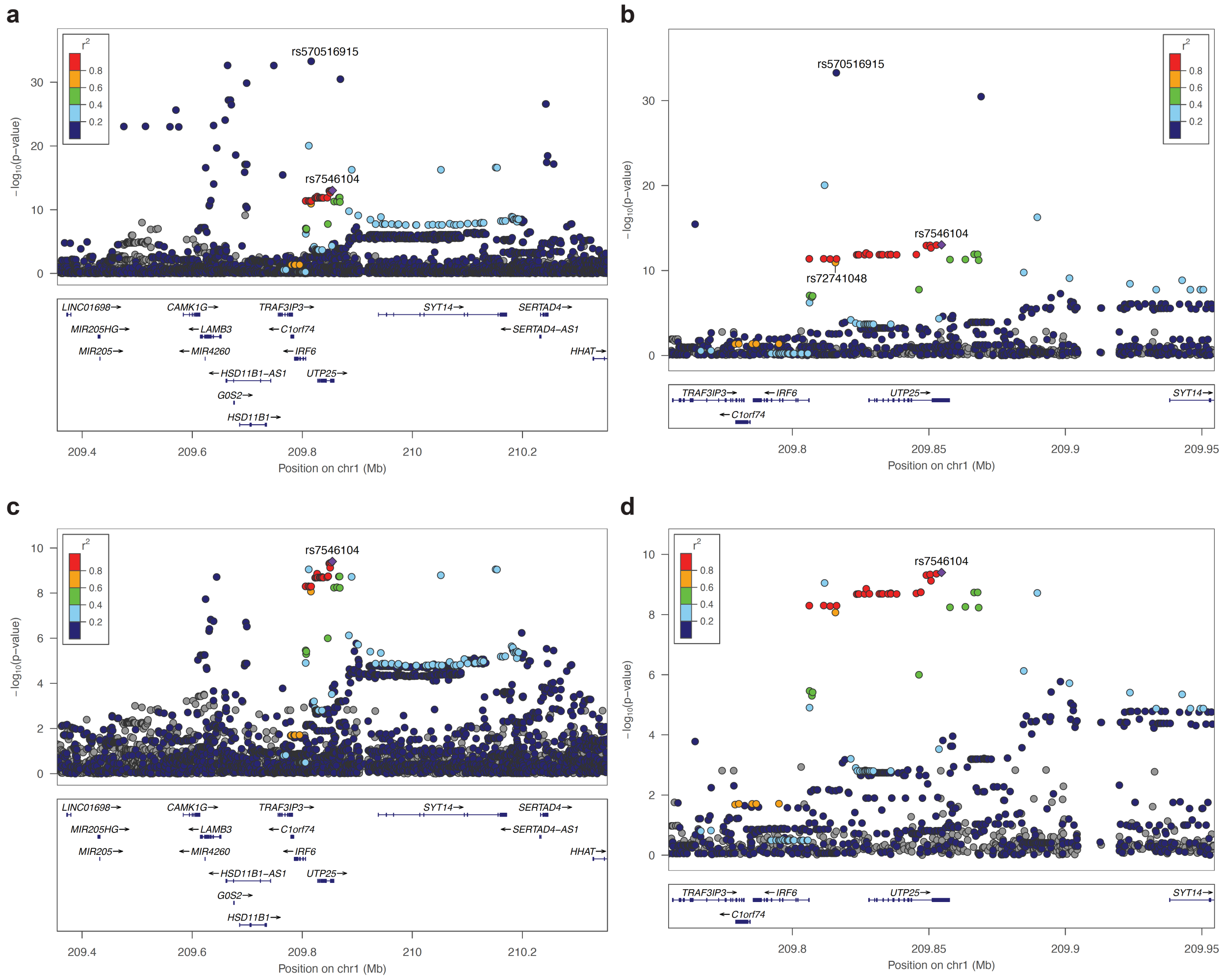


**Supplementary Fig. 5: Regional association plots of the chromosome 1q32.2 locus before (a and b) and after (c and d) conditional analysis.** For clarity, negative $\log_{10} P$ values are shown for variants within a 1 mb (a and c) and a 200 kb (b and d) windows flanking the reference SNP rs7546104, respectively. The reference SNP rs7546104 is labeled with a purple diamond, and pairwise LD ($r^2$) between the reference SNP and other variants are indicated by color. The $r^2$ values were estimated from high-coverage whole-genome sequences of 3,775 Finns. Both directly genotyped and imputed SNPs are plotted. Genomic coordinates are shown according to the human genome build GRCh38/hg38. Plots c and d display SNPs in LD with the lead SNP rs7546104, after adjusting for rs570516915 in a conditional analysis. Plots a and c show 1 mb regions surrounding the index SNP, whereas plots b and d show a zoomed view of a 200 kb region focusing on the independent risk haplotype tagged by rs7546104. SNP rs72741048 has been associated with OFC in a large GWAS in the Chinese population.[1]

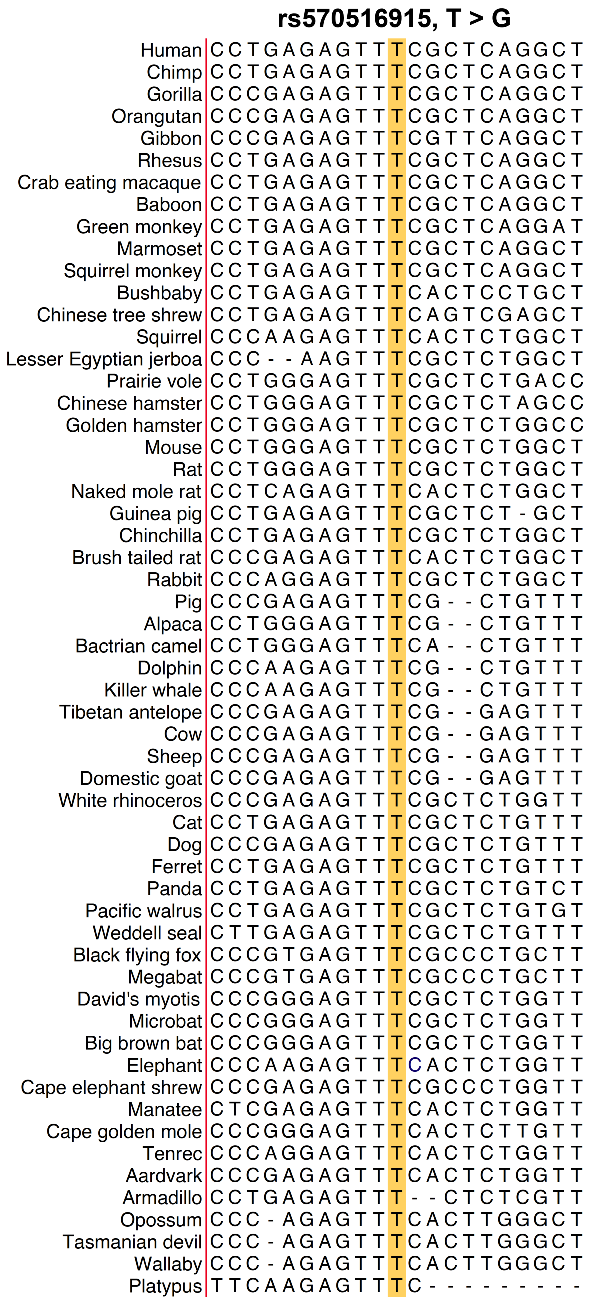


**Supplementary Fig. 6: Evolutionary conservation of the rs570516915 variant site in MCS-9.7.** Multi-species sequence alignments surrounding the rs570516915 variant site in the MCS-9.7 enhancer for *IRF6* were obtained from the multiz alignment and conservation track of 100 vertebrate species in the UCSC Genome Browser (https://genome.ucsc.edu).

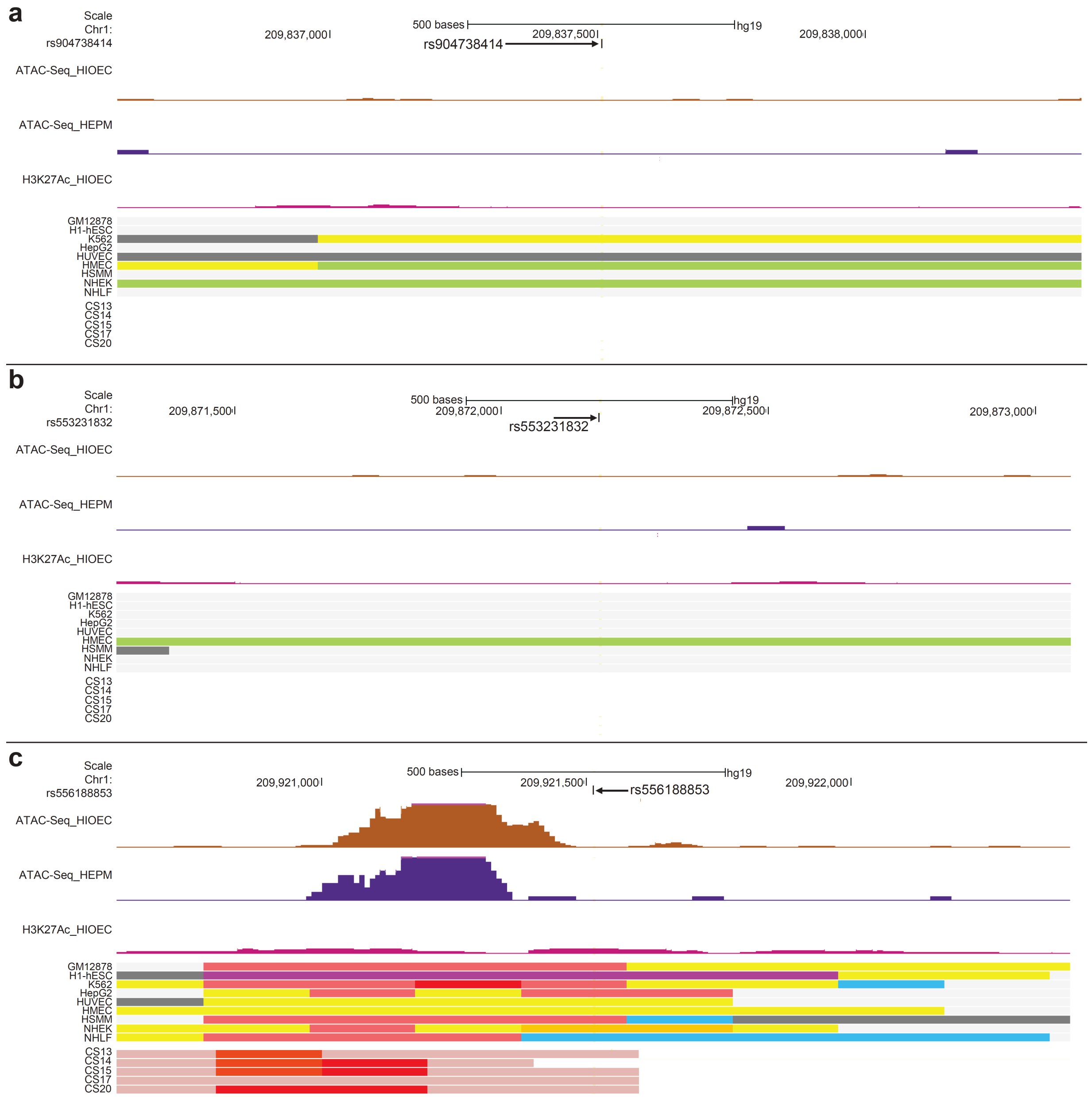


**Supplementary Fig. 7 (a-c): Browser views of the human genome in GRCh37/hg19, illustrating that one (rs556188853) of the 6 SNPs in strong LD with rs570516915 falls into a region with chromatin marks consistent with enhancer activity in epithelial cells (HIOEC and NHEK).** First track, SNPs in LD with rs570516915, (a, rs904738414; b, rs553231832; c, rs556188853;); second and third tracks, ATAC-Seq from HIOEC and HEPM cells, respectively; fourth track, H3K27Ac from HIOEC cells; fifth track, chromatin status revealed by ChIP-Seq to various chromatin marks from the ENCODE Project cell lines and facial explants from human embryos at Carnegie stage (CS) 13-20, color coded as in Fig. 4a**.** Additional color codes from ENCODE project cell lines: bright red, active promoter; light red, weak promoter; purple, inactive/poised promoter. Additional color codes from facial explants from human embryos at CS 13-20: red, active TSS; orange red, promoter upstream or downstream TSS; light purple, poised promoter.

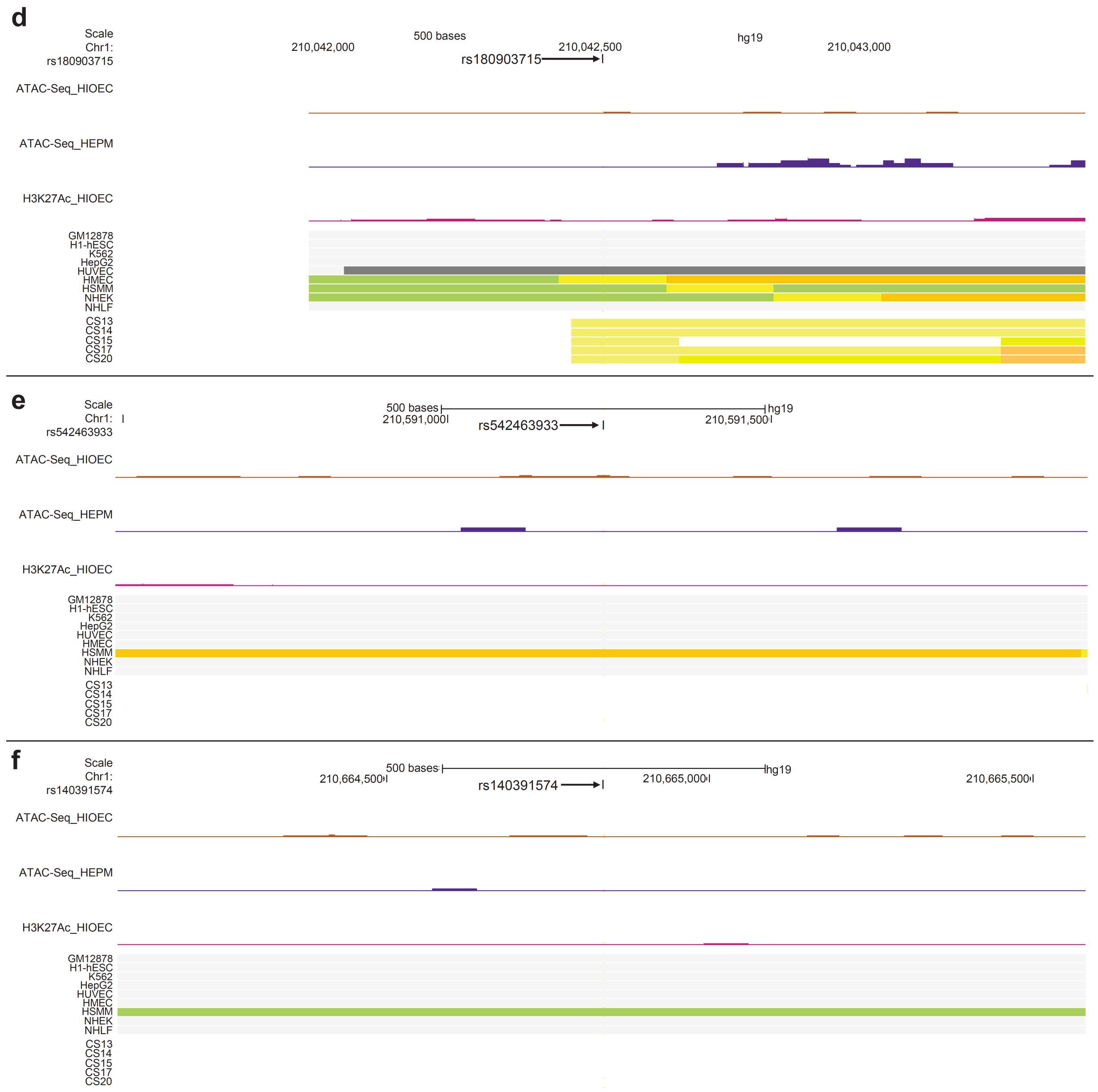


**Supplementary Fig. 7 (d-f): Browser views of the human genome in GRCh37/hg19, illustrating that one (rs556188853) of the 6 SNPs in strong LD with rs570516915 falls into a region with chromatin marks consistent with enhancer activity in epithelial cells (HIOEC and NHEK).** First track, SNPs in LD with rs570516915, (d, rs180903715; e, rs542463933 and f, rs140391574); second and third tracks, ATAC-Seq from HIOEC and HEPM cells, respectively; fourth track, H3K27Ac from HIOEC cells; fifth track, chromatin status revealed by ChIP-Seq to various chromatin marks from the ENCODE Project cell lines and facial explants from human embryos at Carnegie stage (CS) 13-20, color coded as in Fig. 4a.

**a**

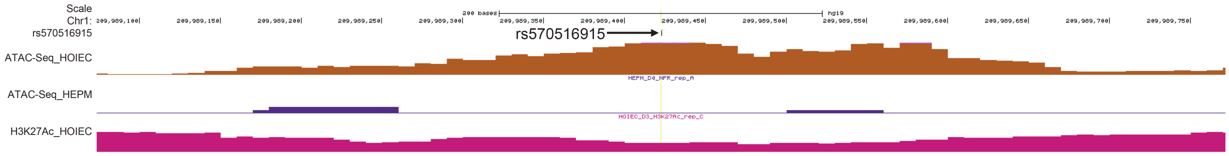


**b**

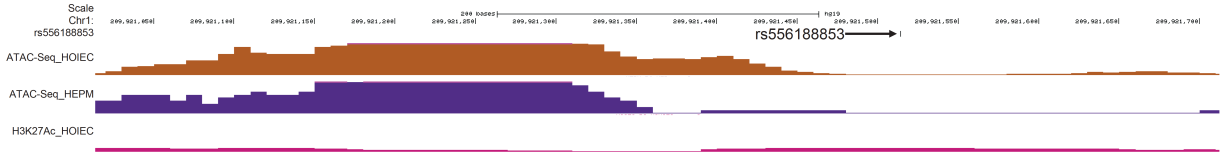


**Supplementary Fig. 8: Browser views of genomic regions included in the luciferase reporter vectors for the rs570516915 and rs556188853 variants**. First track, position of SNPs a) rs570516915 and b) rs556188853; second and third track, ATAC-Seq from HIOEC and HEPM cells, respectively; fourth track, H3K27Ac from HIOEC cells.

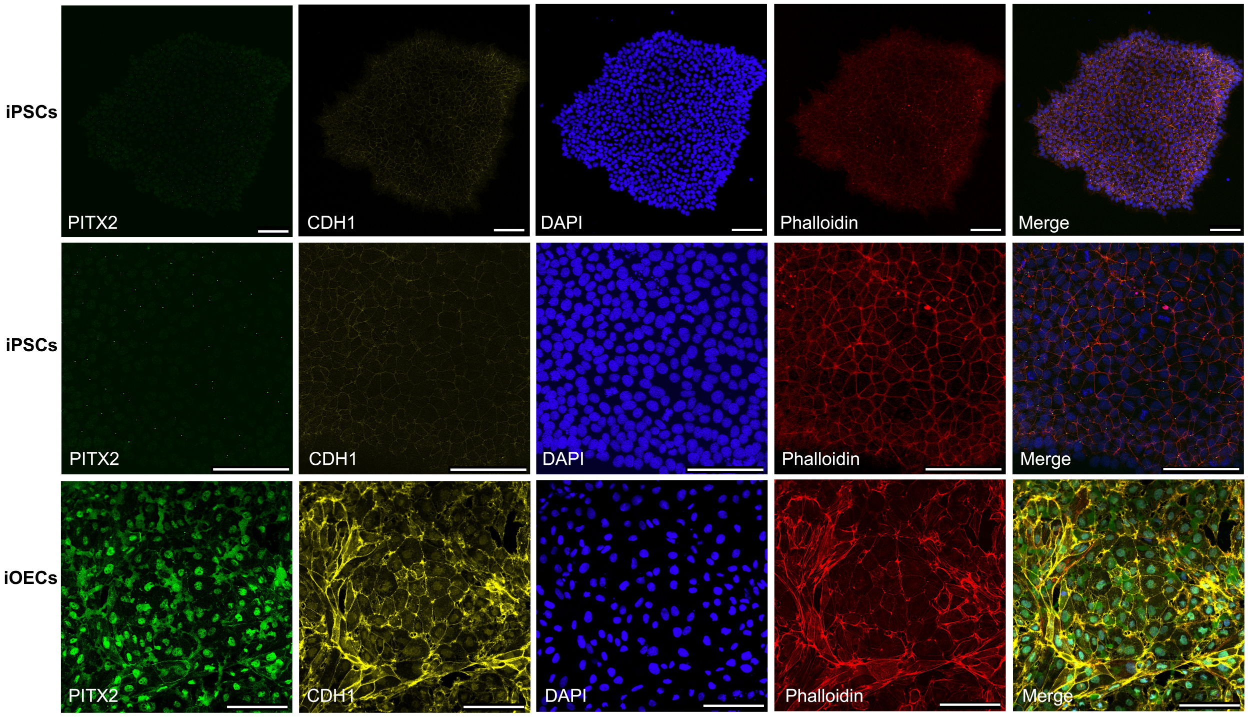


**Supplementary Fig. 9: Immunofluorescence staining of human iPSCs and iOECs.** Green, PITX2-positive nuclei; Yellow, CDH1-positive induced oral epithelial cells (Scale Bar: 100 µm).

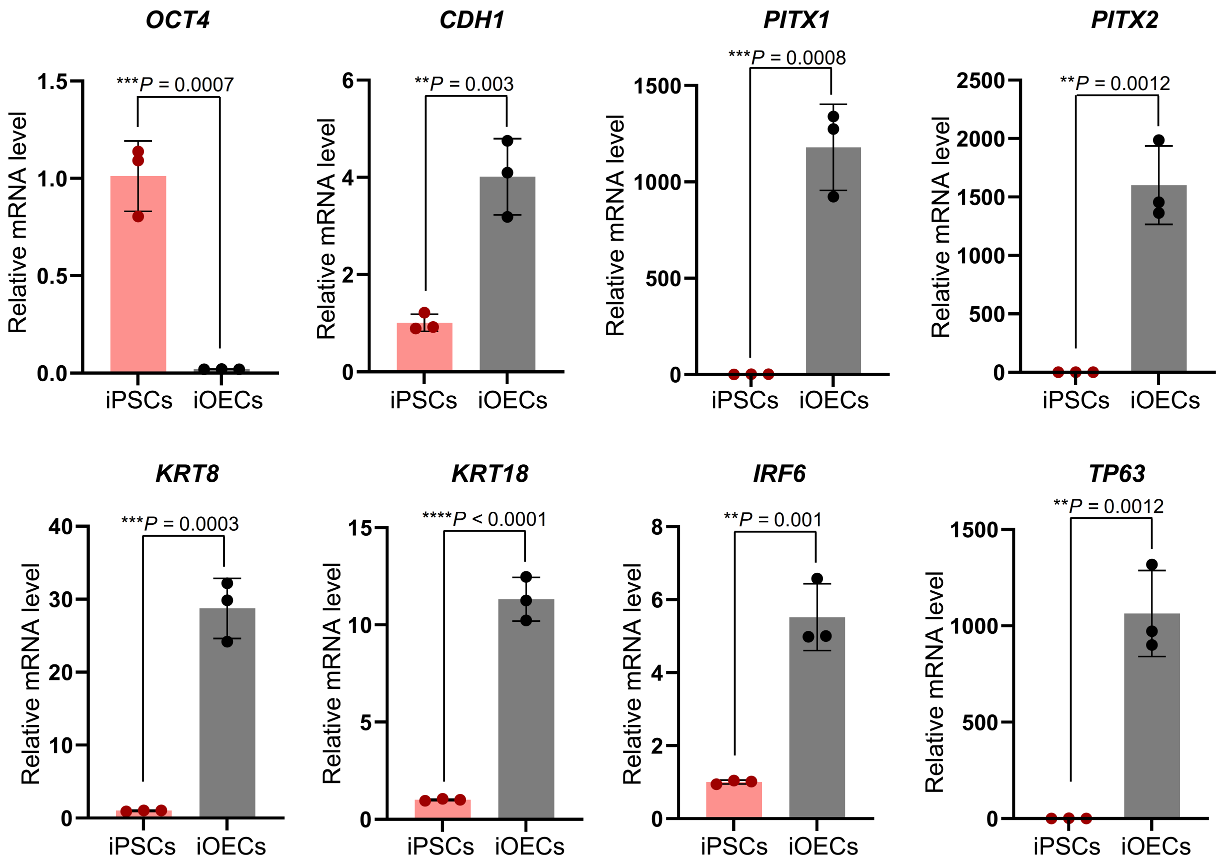


**Supplementary Fig. 10: Scattered dot plots illustrating comparison of expression of stem cell marker (*OCT4*) and different epithelial markers (*CDH1, PITX1, PITX2, KRT8, KRT18*, and *TP63*) in iOECs vs iPSCs by qRT-PCR.** Expression levels of each gene are normalized to *ACTB* (beta actin). Data are represented as mean values +/- s. d. from three replicates. Statistical significance is determined by Student's *t*-test (two-tailed; *P* values are indicated on the plot).

| Rank | Difference log(p) for two sequences | rs570516915_A sequence p-value | rs570516915_C sequence p-value | Matrix_ID | Matrix name |
|---|---|---|---|---|---|
| 1 | 1.8 | 0.00382 | 0.242 | PB0036.1 | Irf6_1 |
| 2 | 1.49 | 0.0117 | 0.36 | PB0034.1 | Irf4_1 |
| 3 | 1.3 | 0.0183 | 0.363 | PB0035.1 | Irf5_1 |
| 4 | -1.13 | 0.354 | 0.026 | PF0167.1 | CCTNTMAGA |
| 5 | 1.02 | 0.0473 | 0.498 | MA0024.1 | E2F1 |
| 6 | 0.906 | 0.111 | 0.898 | MA0393.1 | STE12 |
| 7 | -0.802 | 0.533 | 0.0842 | MA0349.1 | OPI1 |
| 8 | 0.786 | 0.0375 | 0.229 | MA0440.1 | ZAP1 |
| 9 | 0.742 | 0.125 | 0.69 | PB0115.1 | Ehf_2 |
| 10 | 0.737 | 0.156 | 0.852 | MA0360.1 | RDR1 |

**Supplementary Fig. 11: Prediction of IRF6 binding and the effect of the rs570516915 risk allele on its binding affinity.** Sequence shown in Supplementary Fig. 6 was searched in sTRAP[2] (http://trap.molgen.mpg.de/cgi-bin/trap_two_seq_form.cgi) with the non-risk (rs570516915_A) and risk (rs570516915_C) alleles of rs570516915 (AGCCTGAGCG[A/C]AACTCTCAGG). Complementary nucleotides are shown. *P* values are calculated by comparing the observed transcription factor (TF) affinity to the background, using a generalized extreme value (GEV) parametrization model as described previously.[3] *P* values are not corrected for multiple testing. As a visual aid, *P* values less than 0.05 are highlighted in green. TF matrix was selected from Jaspar (all_matrices) and human_promoters was selected as a background model.

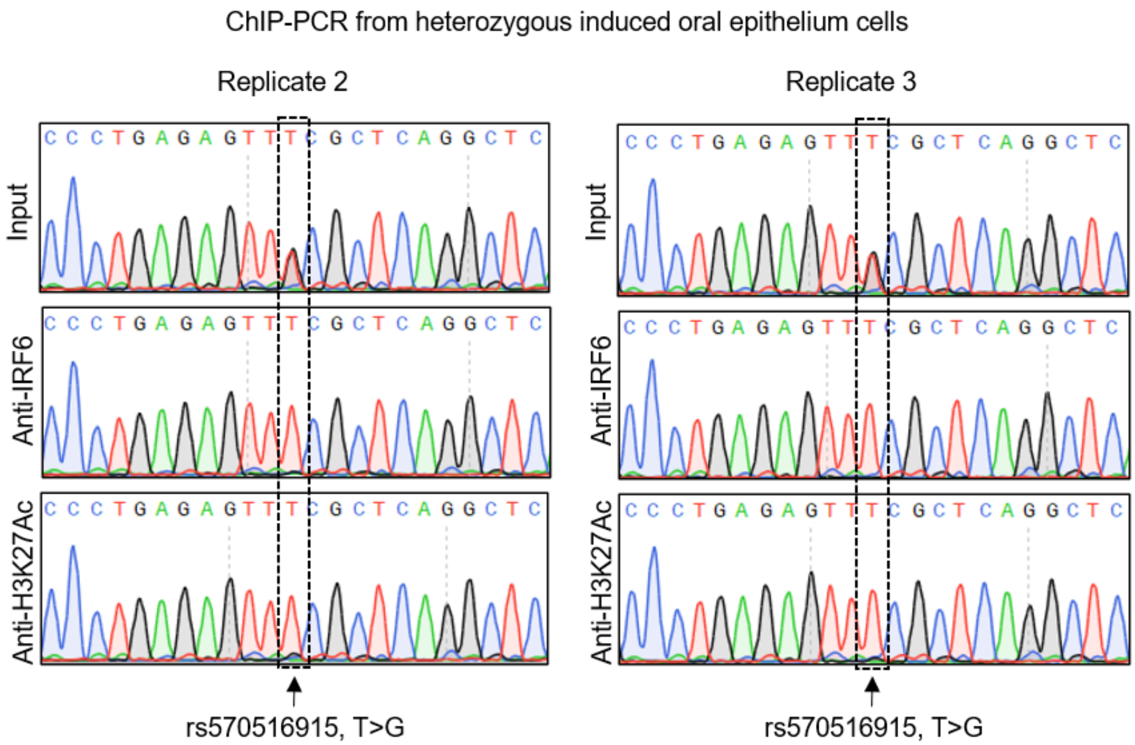


**Supplementary Fig. 12:** Sequencing of anti-IRF6 and anti-H3K27Ac ChIP-PCR product of cells heterozygous for rs570516915 from two ChIP replicates.

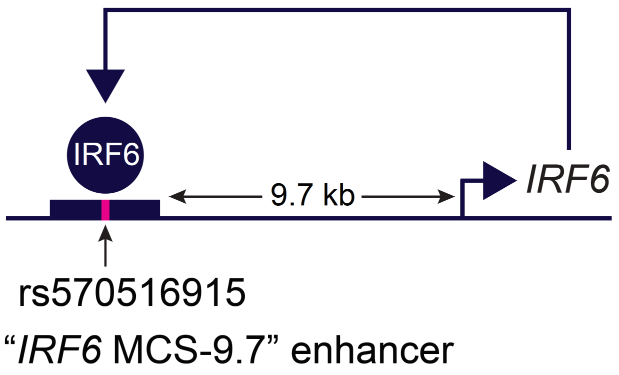


**Supplementary Fig. 13:** A schematic model depicting autoregulation of *IRF6* through the MCS-9.7 enhancer that is 9.7 kb upstream o *IRF6* transcription start site.

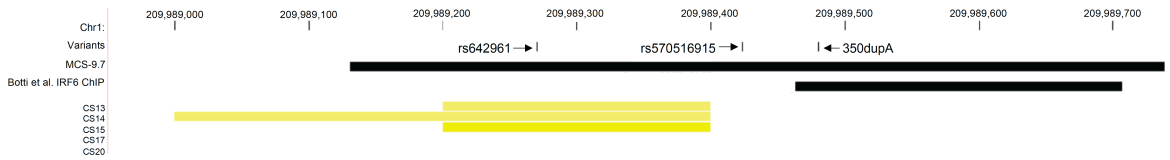


**Supplementary Fig. 14: Browser view of the human genome in GRCh37/hg19, illustrating that rs570516915 is close to IRF6 ChIP-Seq peak identified by Botti et al**. First track,  three DNA variants in MCS-9.7, rs642961[4], rs570516915 and 350dupA[5]; second and third tracks, MCS-9.7[4] and IRF6 binding peaks identified by Botti et al.[6]; fourth track, chromatin status from facial explants from human embryos at Carnegie stage (CS) 13-20, encompassing the time when palate shelves fuse where yellow bars represent the active enhancer elements.

### Supplementary Notes

#### Sequence of the rs556188853 variant region inserted into the luciferase reporter construct. Variant sequence is highlighted in red.

**> hg19_chr1:209921014-209921714**

```
CAGGGGCTCCCTCACACCGTCCTTGAACAGTACCGTGGACCACAAAAGAGCAAGGCTGTCCTAAGGAAAGACTTTGC
CTCTTCGCCGGCCTCCCTTTGGGCCGTCAGGGTGGCAGCGCCACCCCGTGGCTGAATGTGGTGGTGTACCACGCTTC
ACGCAATCTCCTTGCCCAGTGCGTATCAGGTTCTACCCCAGGGCGGCCACCACCTGCACACCAGCGTGCCTACCAGT
GAGTCATCGCTTAGGGCCCTGAACTGACCTAGAGGCGGGGCTGGACTCCGGCGTCCGGTAACTCCGCCCGGCTGCTG
GGGCCTACGTTTGGCATTGAGCCGTGGGGAACTAGACCTGTTCAATAGCGACAGCTAGTGGTCACCCTCAAAAACAA
GGTGCTCTGAACCTCGCGGGGAAAGCGCTGTCAAGATATGGGAAAATCAGGACAGATGATTGGCCTTGAGTGGAAGC
TGGCACATTTTCGAAGCCACTCTTGGGTAAAATGGTCTG/ATCGAAAACCCAAACGCTTTAAAGTGATGTACAACGA
CCTTGGCTGCCTCTCCAGCCACATCTCGGGCCACCTGCCTTGCTTTTGATTCCCCAAGGACAAAGAACTGCCAGTCC
TCTGAAGCACCCTGATGATCCCACTGTGTGTTGGCCATGCCATCTCCTCTGCCCTGAGCGCTCTTCACTCTCCCCAT
TGCATGGTTA
```

#### Sequence of the rs570516915 variant region inserted into the luciferase reporter construct. Variant sequence is highlighted in red.

**> hg19_chr1:209989073-209989773**

```
GTTCATCAGGGGATTTTTTAAAAATTTAGTTTTTAAGAGTTTTATCCAAGGTCTCAAAGCTGGTAAATGGTGAGTAG
GAAGTTGATTCTGCCCGATCTGTCTGACTCCATGCCCTTTCTATTAGGTCATGAAGGGGAACCTGAGGATTGGAGCT
TTGGAATGTTAATCTTACCCAAAGGCCTGAAGTAATACCCCAGAATGTGAACATGTGTGACCATCTGCCTGTCCTGG
GGGTGGGAAGAAGGCAGCATGCTCTATCCTTGACCCTGATTGAGCCCAGGGGCTGAATCTGGAGCTTTGGGGCCTGG
GAACCTCTCTACCTGCGTCAATGTCTGGAGGCCCTGAGAGTTT/GCGCTCAGGCTCAGAGCAGGCATCGCAACCTCC
CAGTTACTATTCTGTGCTGTGGCAAGTGCCAGCTTGTCCTCTCTTCCCCACCCAGCCCGGGAAACCGGCAGCATTTC
TAGTTCAGGCCCAGACCCGTCCTGGCAGCCTGGATTCCACTGCCTAGGCAGGAAGCTCATCTCAGCCCAGTGACCTT
TTCTCTCTGTTTTTTGTCACAGAGGAATTTCCATGCCAGCAGTATGGGGCAATGGGGGTGGGTGGCCAAAGGTTTCC
CCCTTAAGCCACAAGAGCCATGGAGTGGAGGTAAGCTAAGCAAACAGAGGAGGAAGGATGGGAGGGAAGGATCAGGA
AGATTTAGAG
```

#### Sequence of the rs570516915 variant region on the Finnish haplotype background fused to the *LacZ* reporter gene for transgenic F0 embryo assay. Variant sequence is highlighted in red.

**> hg19_chr1:209988937-209990041**

```
CTAAGAGTAATTTACCTATGTTAGCTTTTCTGGAATTGTTCCAGAATCTTGCGCTTTGAAGGAAAAGTTGTGGCTGC
GTATTCTGCCCCTCTCTGTTTGGAAATCCCCTGCACAGCTTTGCCAGCTACTCAGCTTGGTTCATCAGGGGATTTTT
TTAAAATTTAGTTTTTAAGAGTTTTATCCAAGGTCTCAAAGCTGGTAAATGGTGAGTAGGAAGTTGATTCTGCCCGA
TCTGTCTGACTCCATGCCCTTTCTATTAGGTCATGAAGGGGAACCTGAGGATTGGAGCTTTGGAATGTTAATCTTAC
CCAAAGGCCTGAAGTAATACCCCAGGATGTGAACATGTGTGACCATCTGCCTGTCCTGGGGGTGGGAAGAAGGCAGC
ATGCTCTATCCTTGACCCTGATTGAGCCCAGGGGCTGAATCTGGAGCTTTGGGGCCTGGGAACCTCTCTACCTGCGT
CAATGTCTGGAGGCCCTGAGAGTTT/GCGCTCAGGCTCAGAGCAGGCATCGCAACCTCCCAGTTACTATTCTGTGCT
GTGGCAAGTGCCAGCTTGTCCTCTCTTCCCCACCCAGCCCGGGAAACCAGCAGCATTTCTAGTTCAGGCCCAGACCC
GTCCTGGCAGCCTGGATTCCACTGCCTAGGCAGGAAGCTCATCTCAGCCCAGTGACCTTTTCTCTCTGTTTTTTGTC
ACAGAGGAATTTCCATGCCAGCAGTATGGGGCAATGGGGGTGGGTGGCCAAAGGTTTCCCCCTTAAGCCACAAGAGC
CATGGAGTGGAGGTAAGCTAAGCAAACAGAGGAGGAAGGATGGGAGGGAAGGATCAGGAAGATTTAGAGAGTCCATT
CCTCAGGCTGCTTCATCCTCAAATTCCAAGGTAAATAAAGGTGTTGGGAAGGATGCACTATATGTCCAGCTGCCCAG
CCCTGAAGATCCTCCCCCTAGAGAACTGAGACAGGAGTTTTCTCACATTCCCACATGCAGGAGGGAAAGAGGCTGGG
CCTGGGCAGGTCTAGAGAGTTCTGTTCCCCTGCCCCAGGGGAACACGCCTAGGCTTGCTGACCTCTCAGGTAGGAGT
CTGTCAGGTTGGAAGCTGGGCCTCTCATT
```
